# An international multicenter examination of MOG antibody assays

**DOI:** 10.1101/19011049

**Authors:** Markus Reindl, Kathrin Schanda, Mark Woodhall, Fiona Tea, Sudarshini Ramanathan, Jessica Sagen, Jim Fryer, John Mills, Bianca Teegen, Swantje Mindorf, Nora Ritter, Ulrike Krummrei, Winfried Stöcker, Juliane Eggert, Eoin P. Flanagan, Melanie Ramberger, Harald Hegen, Kevin Rostasy, Thomas Berger, Maria Isabel Leite, Jacqueline Palace, Sarosh R. Irani, Russell C. Dale, Christian Probst, Monika Probst, Fabienne Brilot, Sean Pittock, Patrick Waters

## Abstract

**Objectives:** To compare the reproducibility of 11 antibody assays for IgG and IgM myelin oligodendrocyte glycoprotein antibodies (MOG-IgG, MOG-IgM) from five international centers.

**Methods:** The following samples were analyzed: MOG-IgG clearly positive sera (n=39), MOG-IgG low positive sera (n=39), borderline negative sera (n=13), clearly negative sera (n=40), and healthy blood donors (n=30). As technical controls, 18 replicates (9 MOG-IgG positive and 9 negative) were included. All samples and controls were re-coded, aliquoted, and distributed to the five testing centers which performed the following antibody assays: five live and one fixed immunofluorescence cell-based assays (CBA-IF, five MOG-IgG, one MOG-IgM), three live flow cytometry cell-based assays (FACS-CBA, all MOG-IgG), and two enzyme-linked immunosorbent assays (ELISA, both MOG-IgG).

**Results:** We found excellent agreement (96%) between the live CBAs for MOG-IgG for samples previously identified as clearly positive or negative from four different national testing centers. The agreement was lower with fixed CBA-IF (90%) and the ELISA showed no concordance with CBAs for detection of human MOG-IgG. All CBAs showed excellent inter-assay reproducibility. The agreement of MOG-IgG CBAs for borderline negative (77%) and particularly low positive (33%) samples was less good. Finally, most samples from healthy blood donors (97%) were negative for MOG-IgG in all CBAs.

**Conclusion:** Live MOG-IgG CBAs showed excellent agreement for high positive and very good agreement for negative samples at four international testing centers. Low positive samples were more frequently discordant than in similar assays for other autoantigens. Further research is needed to improve international standardization for clinical care.

## INTRODUCTION

Immunoglobulin G antibodies to myelin-oligodendrocyte-glycoprotein (MOG-IgG) are found in adults and children who present with a spectrum of central nervous system features that include optic neuritis, acute disseminated encephalomyelitis (ADEM), myelitis, seizures, encephalitis, brainstem and/or cerebellar involvement. In addition, the presence of MOG-IgG can discriminate these disorders from multiple sclerosis (MS) ^1^. Numerous studies have used different immunoassays for MOG-IgG detection, but it is now clear that native full-length human MOG as an assay substrate is crucial to make this clinical distinction. When measured using older generation assays (ELISA, western blot), MOG-IgG are prevalent and have been identified in healthy individuals and patients with a wide variety of clinical presentations. Thus, their detection was initially considered to have little clinical utility. However, when measured by live cell-based assays (CBA) an association between MOG-IgG antibodies and a non-MS demyelinating phenotype has been established. This understanding has driven the establishment of different variants of MOG-IgG assays with native MOG substrates in multiple centers world-wide. There is limited data on assay reproducibility between these centers. In this study, we compared the most frequently used assays for MOG-IgG detection, such as live and fixed immunofluorescence cell-based assays (CBA-IF) ^2-18^, live flow cytometry cell-based assays (CBA-FACS) ^4, 19-28^, and enzyme-linked immunosorbent assays (ELISA) ^29, 30^.

## METHODS

### Patients and controls

The clinical laboratories (Innsbruck, Mayo Clinic, Oxford, and Sydney; centers 1-4) sent the following groups of coded serum samples and clinical information to the Institute for Quality Assurance (IfQ; Lübeck, Germany):

#### Phase I: 89 coded samples sent to centers 1-4 and center 5 (Euroimmun) for testing (figure 1)

1. MOG-IgG clearly positive: thirty-nine blinded samples from all labs with a previously determined clearly positive MOG-Ab serostatus (high titers or FACS binding ratios, supplementary methods, etable 2), all of them diagnosed with inflammatory demyelinating diseases known to be associated with MOG-IgG (such as ADEM, AQP4-antibody negative neuromyelitis optica spectrum disorder (NMOSD), optic neuritis, myelitis, and other demyelinating diseases).

**Figure 1:**
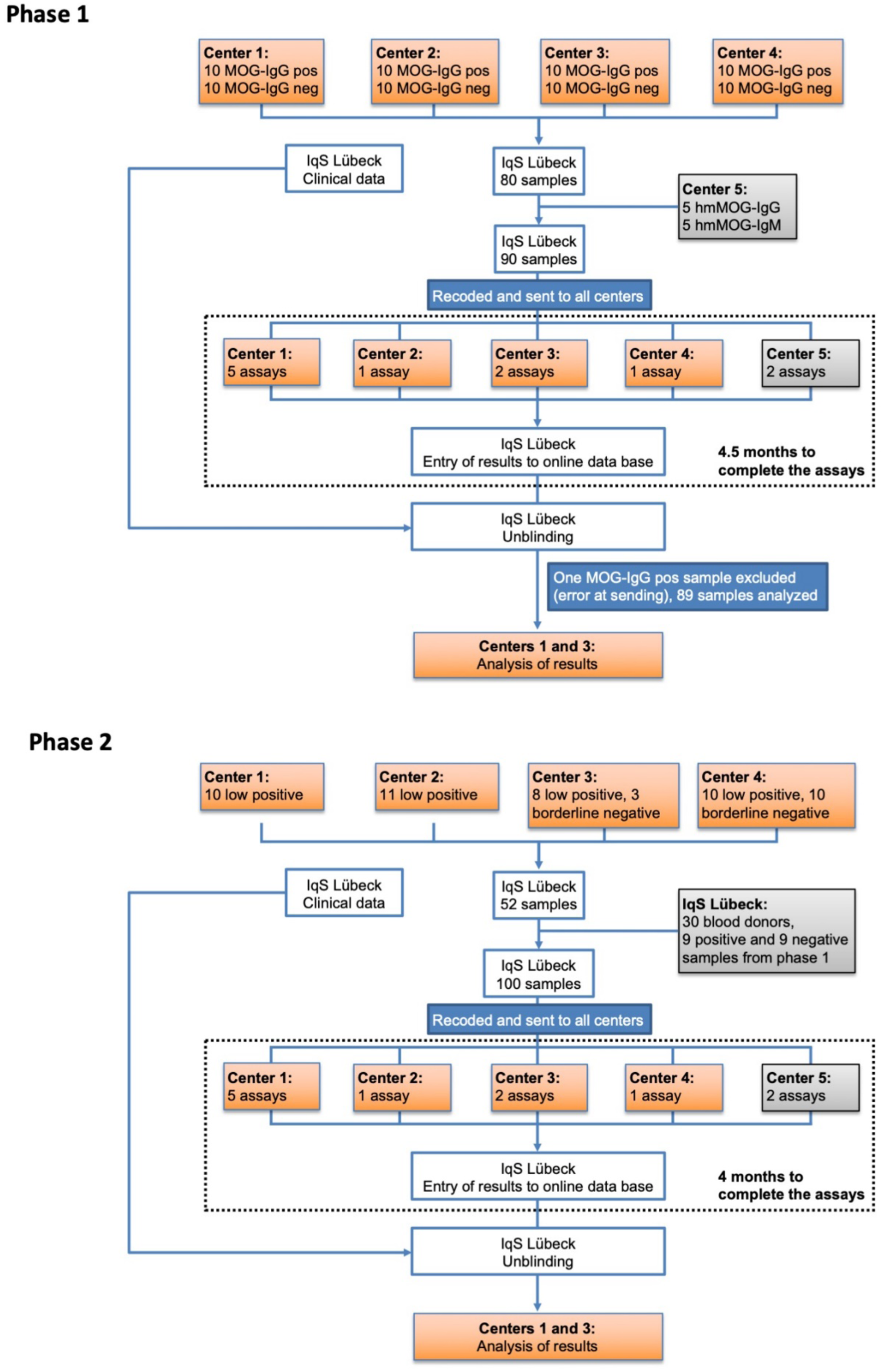
Flow chart showing phases I and II of this study. Center 1 (Innsbruck) performed 5 assays (live CBA-IF MOG-IgG(H+L), live CBA-IF MOG-IgG(Fc), live CBA-FACS MOG-IgG(Fc), live CBA-IF MOG-IgM and ELISA MOG-IgG); center 2 (Mayo Clinic) performed one assay (live CBA-FACS MOG-IgG1); center 3 (Oxford) performed 2 assays (live CBA-IF MOG-IgG(H+L) and live CBA-IF MOG-IgG1); center 4 (Sydney) performed one assay (live CBA-FACS MOG-IgG(H+L)) which was repeated twice; center 5 (Euroimmun) performed 2 assays (fixed CBA-IF MOG-IgG(Fc) and ELISA MOG-IgG(Fc)).
2. MOG-IgG clearly negative (negative or very low titers or FACS binding ratios, supplementary methods, etable 2): forty blinded samples from all labs with a previously determined clearly negative MOG-Ab serostatus. Eighteen of the forty samples were from people who also presented with clinically overlapping features such as optic neuritis, myelitis, ADEM, or encephalitis. The other samples were from controls (seven from people with MS, five from people with other neurological diseases and ten from healthy controls).
3. Ten technical controls (humanized monoclonal MOG-Ab 8-18-C5 ^31^, five samples IgG1 and five samples IgM (kappa) in different dilutions, but of unknown IgG or IgM concentration, contributed by center 5.

#### Phase II: 100 coded samples sent to five centers for testing (18 repeat and 82 new, figure 1)

4. Nine positive and nine negative samples from phase I were sent out a second time to assess inter-assay variations.
5. Thirty healthy blood donors were contributed by the IfQ. No clinical information was available and samples were not pretested for antibodies against MOG or other autoantigens.
6. MOG-IgG low/borderline positive: thirty-nine blinded samples from all labs with a previously determined low positive MOG-IgG serostatus (just above the individual cut-off values, supplementary methods, etable 2). Thirty-six of these samples were from people with inflammatory demyelinating diseases associated with MOG-IgG and three were from MS patients.
7. MOG-IgG borderline negative: thirteen blinded samples from all labs with a previously determined borderline negative MOG-IgG serostatus (just below the individual cut-off values, supplementary methods, etable 2). Five of these samples were from patients with inflammatory demyelinating diseases associated with MOG-IgG and eight were from controls (three from people with MS and five from people with other neurological diseases).

### Standard protocol approvals, registrations, and patient consents

The present study was approved by the ethical committees of Medical University of Innsbruck (AM3041A and AM4059), Oxford (REC 16/SC/0224), Mayo Clinic (IRB 08-007810) and Sydney (NEAF 12/SCHN/395). All samples were anonymized before sending to the blinding center.

### Laboratory methods and analysis

All samples and controls were re-coded, aliquoted, and distributed by an investigator not involved in antibody testing from the IfQ, Lübeck, Germany, to the five testing centers which performed the 7 live CBAs (4 CBA-IF and 3 CBA-FACS) one fixed CBA-IF for MOG-IgG, one live CBA-IF for MOG-IgM and 2 ELISAs for MOG-IgG in the two study phases (figure 1, table 1 and supplementary methods).

**Table 1.**
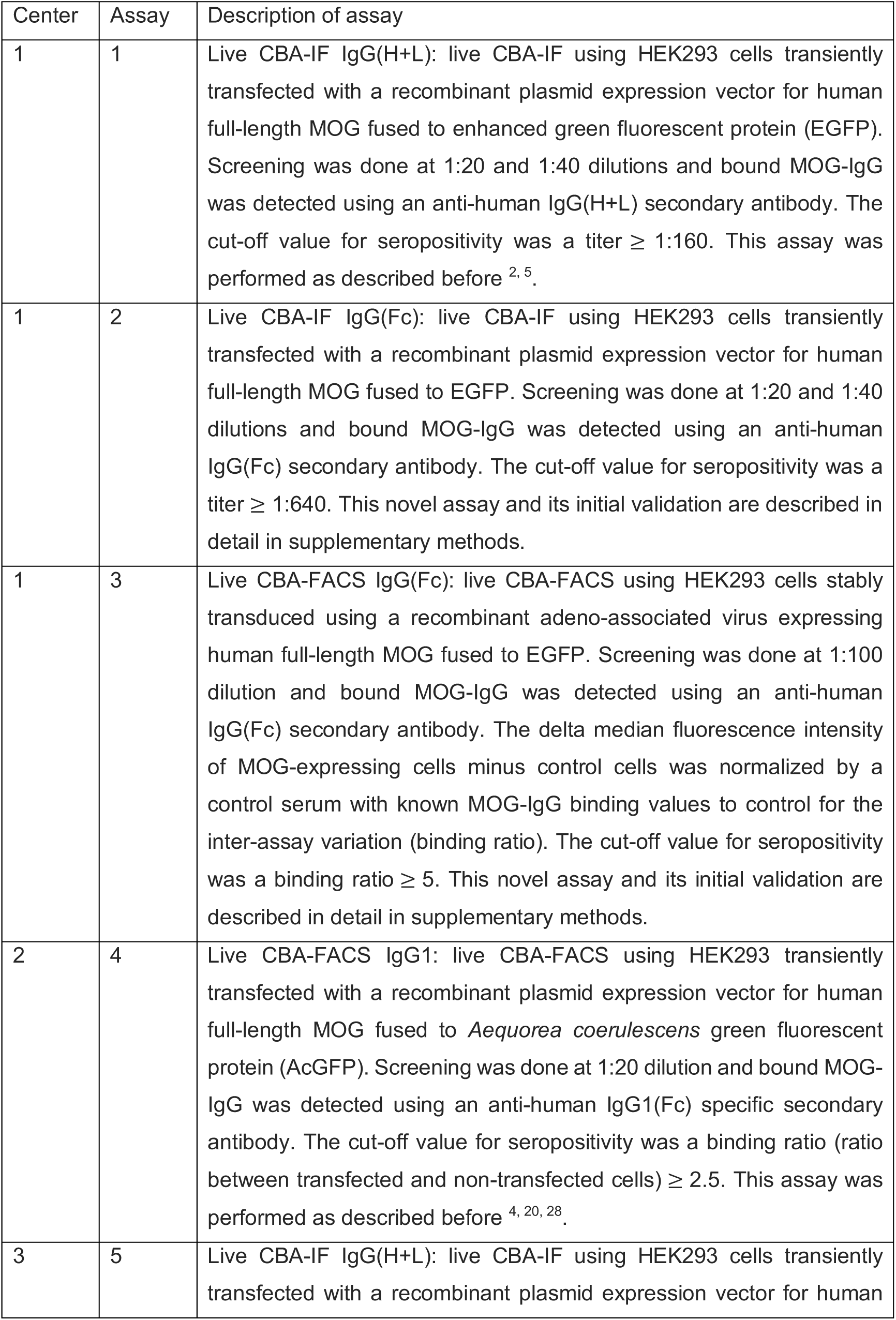
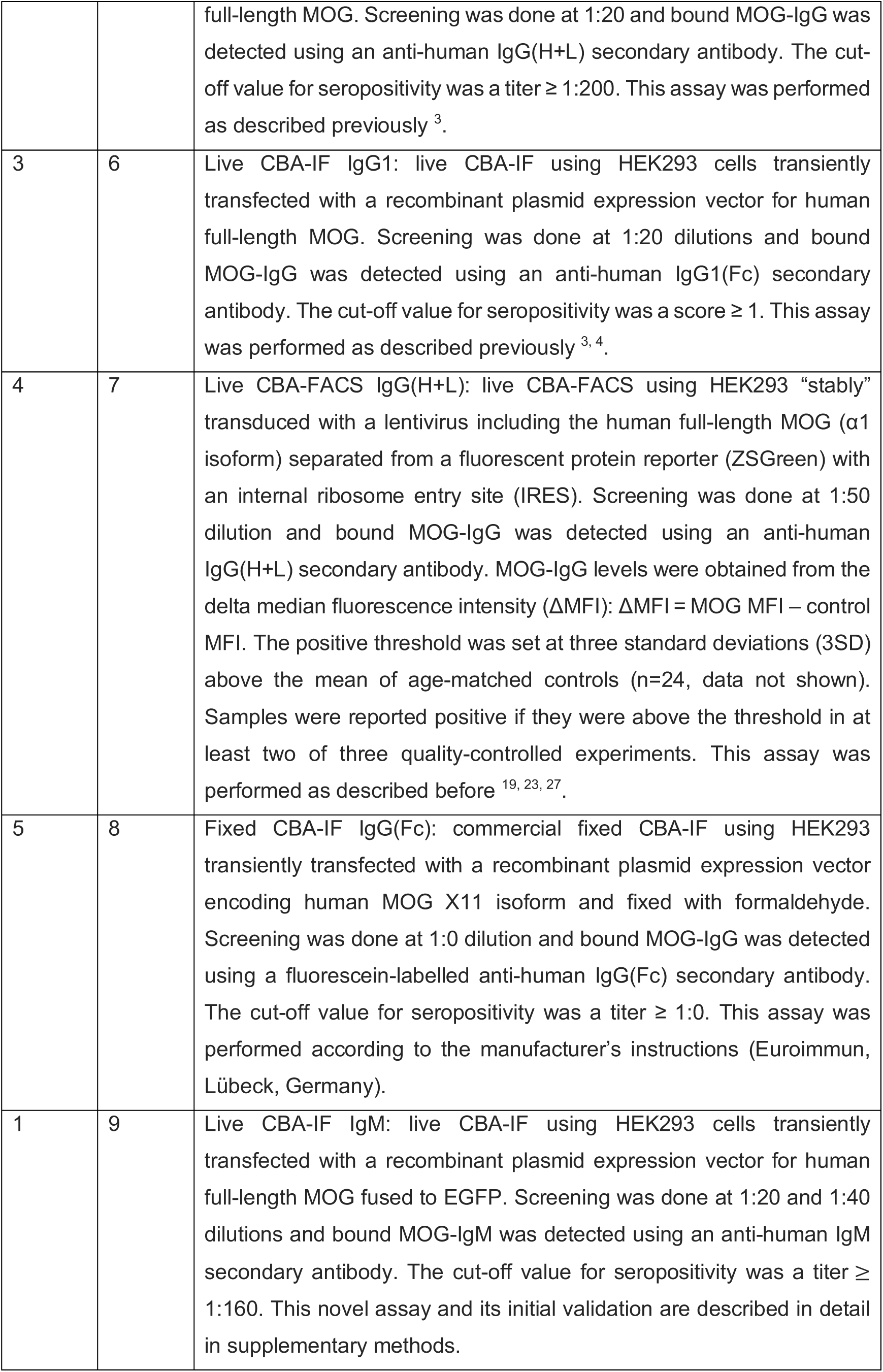
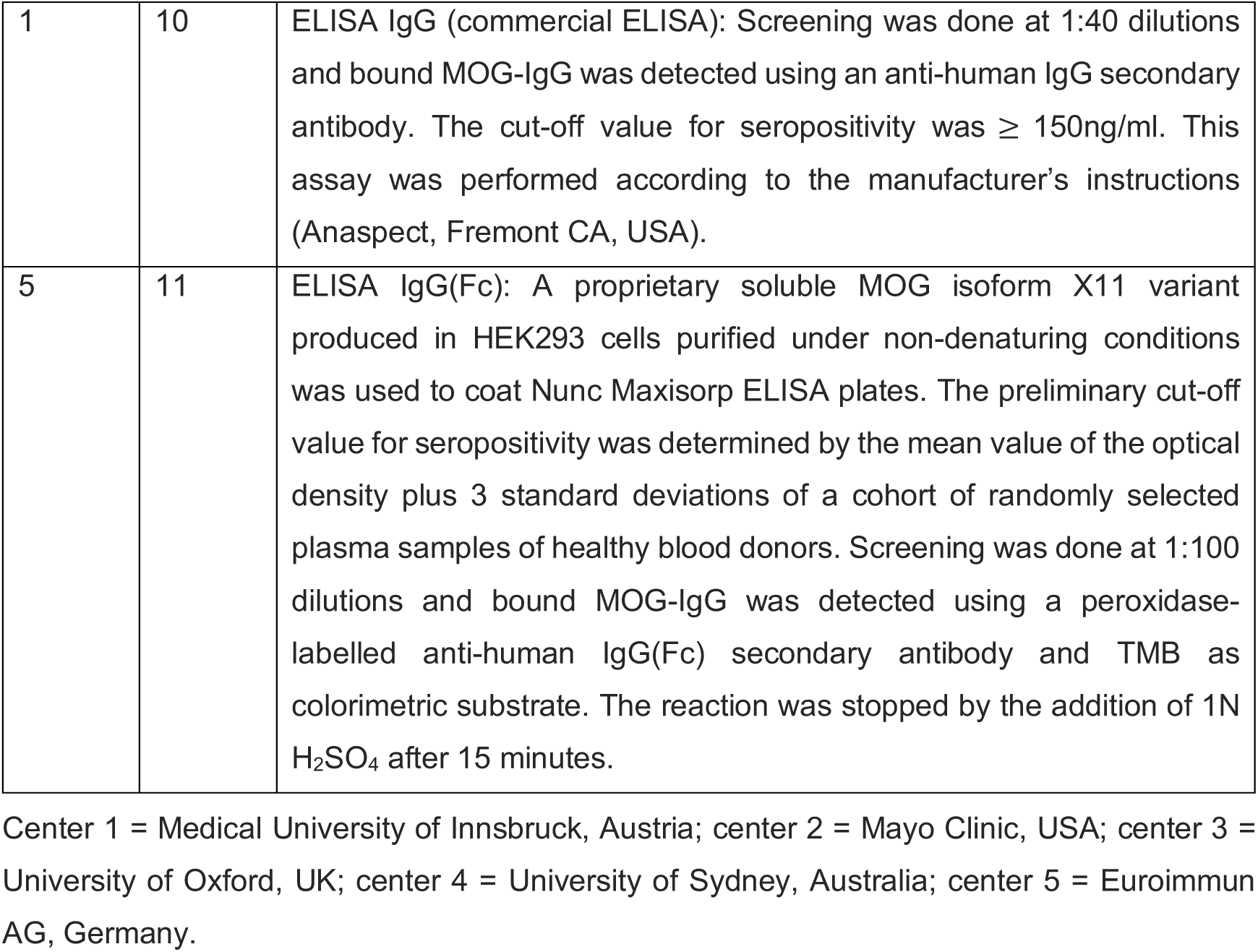
Description of immunoassays used for the measurement of MOG-IgG and MOG-IgM antibodies.

| Center | Assay | Description of assay |
| --- | --- | --- |
| 1 | 1 | Live CBA-IF IgG(H+L): live CBA-IF using HEK293 cells transiently transfected with a recombinant plasmid expression vector for human full-length MOG fused to enhanced green fluorescent protein (EGFP). Screening was done at 1:20 and 1:40 dilutions and bound MOG-IgG was detected using an anti-human IgG(H+L) secondary antibody. The cut-off value for seropositivity was a titer $\geq 1:160$. This assay was performed as described before <sup>2, 5</sup>. |
| 1 | 2 | Live CBA-IF IgG(Fc): live CBA-IF using HEK293 cells transiently transfected with a recombinant plasmid expression vector for human full-length MOG fused to EGFP. Screening was done at 1:20 and 1:40 dilutions and bound MOG-IgG was detected using an anti-human IgG(Fc) secondary antibody. The cut-off value for seropositivity was a titer $\geq 1:640$. This novel assay and its initial validation are described in detail in supplementary methods. |
| 1 | 3 | Live CBA-FACS IgG(Fc): live CBA-FACS using HEK293 cells stably transduced using a recombinant adeno-associated virus expressing human full-length MOG fused to EGFP. Screening was done at 1:100 dilution and bound MOG-IgG was detected using an anti-human IgG(Fc) secondary antibody. The delta median fluorescence intensity of MOG-expressing cells minus control cells was normalized by a control serum with known MOG-IgG binding values to control for the inter-assay variation (binding ratio). The cut-off value for seropositivity was a binding ratio $\geq 5$. This novel assay and its initial validation are described in detail in supplementary methods. |
| 2 | 4 | Live CBA-FACS IgG1: live CBA-FACS using HEK293 transiently transfected with a recombinant plasmid expression vector for human full-length MOG fused to *Aequorea coerulescens* green fluorescent protein (AcGFP). Screening was done at 1:20 dilution and bound MOG-IgG was detected using an anti-human IgG1(Fc) specific secondary antibody. The cut-off value for seropositivity was a binding ratio (ratio between transfected and non-transfected cells) $\geq 2.5$. This assay was performed as described before <sup>4, 20, 28</sup>. |
| 3 | 5 | Live CBA-IF IgG(H+L): live CBA-IF using HEK293 cells transiently transfected with a recombinant plasmid expression vector for human |
| | | full-length MOG. Screening was done at 1:20 and bound MOG-IgG was detected using an anti-human IgG(H+L) secondary antibody. The cut-off value for seropositivity was a titer $\geq 1:200$. This assay was performed as described previously <sup>3</sup>. |
| 3 | 6 | Live CBA-IF IgG1: live CBA-IF using HEK293 cells transiently transfected with a recombinant plasmid expression vector for human full-length MOG. Screening was done at 1:20 dilutions and bound MOG-IgG was detected using an anti-human IgG1(Fc) secondary antibody. The cut-off value for seropositivity was a score $\geq 1$. This assay was performed as described previously <sup>3, 4</sup>. |
| 4 | 7 | Live CBA-FACS IgG(H+L): live CBA-FACS using HEK293 “stably” transduced with a lentivirus including the human full-length MOG ($\alpha 1$ isoform) separated from a fluorescent protein reporter (ZSGreen) with an internal ribosome entry site (IRES). Screening was done at 1:50 dilution and bound MOG-IgG was detected using an anti-human IgG(H+L) secondary antibody. MOG-IgG levels were obtained from the delta median fluorescence intensity (ΔMFI): ΔMFI = MOG MFI – control MFI. The positive threshold was set at three standard deviations (3SD) above the mean of age-matched controls (n=24, data not shown). Samples were reported positive if they were above the threshold in at least two of three quality-controlled experiments. This assay was performed as described before <sup>19, 23, 27</sup>. |
| 5 | 8 | Fixed CBA-IF IgG(Fc): commercial fixed CBA-IF using HEK293 transiently transfected with a recombinant plasmid expression vector encoding human MOG X11 isoform and fixed with formaldehyde. Screening was done at 1:0 dilution and bound MOG-IgG was detected using a fluorescein-labelled anti-human IgG(Fc) secondary antibody. The cut-off value for seropositivity was a titer $\geq 1:0$. This assay was performed according to the manufacturer’s instructions (Euroimmun, Lübeck, Germany). |
| 1 | 9 | Live CBA-IF IgM: live CBA-IF using HEK293 cells transiently transfected with a recombinant plasmid expression vector for human full-length MOG fused to EGFP. Screening was done at 1:20 and 1:40 dilutions and bound MOG-IgM was detected using an anti-human IgM secondary antibody. The cut-off value for seropositivity was a titer $\geq 1:160$. This novel assay and its initial validation are described in detail in supplementary methods. |
| 1 | 10 | ELISA IgG (commercial ELISA): Screening was done at 1:40 dilutions and bound MOG-IgG was detected using an anti-human IgG secondary antibody. The cut-off value for seropositivity was $\geq 150\text{ng/ml}$. This assay was performed according to the manufacturer's instructions (Anaspect, Fremont CA, USA). |
| 5 | 11 | ELISA IgG(Fc): A proprietary soluble MOG isoform X11 variant produced in HEK293 cells purified under non-denaturing conditions was used to coat Nunc Maxisorp ELISA plates. The preliminary cut-off value for seropositivity was determined by the mean value of the optical density plus 3 standard deviations of a cohort of randomly selected plasma samples of healthy blood donors. Screening was done at 1:100 dilutions and bound MOG-IgG was detected using a peroxidase-labelled anti-human IgG(Fc) secondary antibody and TMB as colorimetric substrate. The reaction was stopped by the addition of 1N H<sub>2</sub>SO<sub>4</sub> after 15 minutes. |

Center 1 = Medical University of Innsbruck, Austria; center 2 = Mayo Clinic, USA; center 3 = University of Oxford, UK; center 4 = University of Sydney, Australia; center 5 = Euroimmun AG, Germany.

### Statistical analysis

Upon completion of the testing, the assay results from each center were entered onto a web-based database. The data were then unblinded and analyzed. Statistical analyses were carried out using IBM SPSS software (release 24.0, IBM, Armonk, NY) or GraphPad Prism 8 (GraphPad, San Diego, CA). Correlation of parameters was analyzed with Spearman’s non-parametric correlation. Kappa statistics was used to assess the concordance between assays. All graphs were created using GraphPad Prism.

### Data availability statement

The dataset used and analyzed during the current study is included in the main text and the supplementary files.

## RESULTS

### CBAs for MOG-IgG show a very good agreement on clear positive and negative samples

In the first phase of this study (figure 1, phase 1) all centers analyzed samples sent as clearly positive (n=39) or negative (n=40) by centers 1-4 (figure 2 A, etable 3). In general, there was a very good agreement for the 8 MOG-IgG CBAs (figure 2 B): 39/40 (97.5%) samples sent as negative were negative in all 8 CBAs. This agreement was 100% if the fixed commercial CBA was excluded. For the samples submitted as positive 32/39 (82%) samples were concordant across all 8 CBAs; again, this improved to 92% (36/39) if the fixed CBA was excluded. Overall there was 96% concordance across all samples when tested on live platforms in four international testing centers. The concordance dropped to 90% if the results of the fixed CBA tested in-house at center 5 were included.

**Figure 2:**
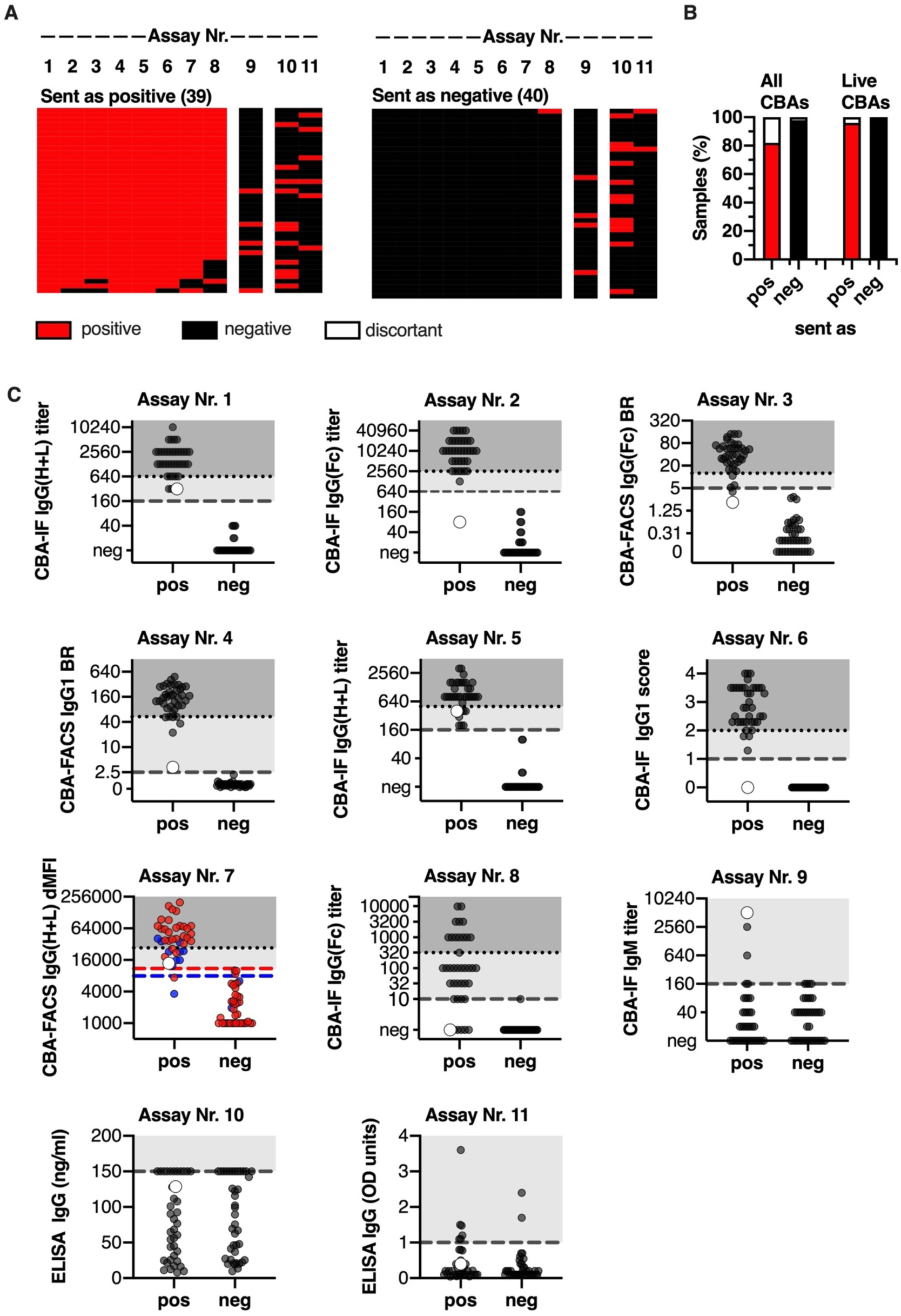
(A) Heatmap of the qualitative results for samples sent as clearly positive (n=39) or negative (n=40). Each column is an individual assay (1-8 MOG-IgG CBAs, 9 MOG-IgM CBA, 10-11 MOG-IgG ELISA) and each row is an individual serum sample. Results are based on qualitative results; negative samples are black and positive samples are red. The samples are shown according to their serostatus sent by the individual centers. (B) Agreement of MOG-IgG CBAs according to the samples sent (pos=positive, neg=negative). Results (in % of all samples) are grouped according to their agreement in all 8 CBAs or in the 7 live CBAs (red: positive in all CBAs, black: negative in all CBAs, white: discordant). (C) Quantitative results for all assays. The cut-off values for all assays except assay Nr. 7 are indicated by the dashed grey lines. For assay Nr. 7 cut-off levels for pediatric samples (blue dots) are indicated by the blue dashed line and cut-off levels for adult samples (red dots) are indicated by the red dashed line. The quantitative range of each assay result for its probability to be seropositive in all live CBAs is indicated by the dotted line and shaded in darker grey (100% probability), whereas the range of discordant samples is shaded in light grey. A single sample with high IgM titer 1:5120 and low positive in the IgG(H+L) and on IgG1, but not in another IgG1 and the IgG(Fc) assays is indicated by the larger white dot. BR=binding ratio, dMFI = delta mean fluorescence intensity.

MOG-IgMs at a titer ≥ 1:160 were a rare finding in samples sent as clear positive (5/39, 13%). One of these 5 samples had a high MOG-IgM titer (1:5120, figure 2 C, assay Nr. 9, large grey dot) and was low positive for MOG-IgG in four assays (using IgG(H+L) and IgG1 secondary antibodies), but negative in 4 assays (using IgG(Fc) and IgG1 secondary antibodies). The other 4 samples were positive for MOG-IgG in all CBAs. MOG-IgMs at a titer ≥ 1:160 were absent in all 40 samples sent as negative.

Overall, there was excellent agreement between the seven live MOG-IgG CBAs (median kappa value 0.975, range 0.924 to 1.000). The agreement of the fixed MOG-IgG CBA with the live MOG-IgG CBAs was very good (median kappa value 0.822, range 0.821 to 0.847). There was no agreement between the MOG-IgG and MOG-IgM CBAs (median kappa value −0.003, range −0.180 to 0.103), or between the MOG-IgG CBAs and the ELISA (median kappa value 0.112, range 0.105 to 0.203).

The quantitative values for all assays are provided in figure 2 C. Most of the assays had a very clear separation of positive and negative samples. The quantitative range in which an individual sample is positive in all live CBAs is indicated by the dotted line and shaded in darker grey (100% probability). For example, if a result of assay Nr. 1 is positive with a MOG-IgG titer of 1:1280 then there is 100% probability that this result is also positive in all other live CBAs. However, on the same assay a MOG-IgG titer of 1:320 (light grey area) is more likely to have discrepant results between centers.

It is evident that there is very good correlation of quantitative results for the seven live MOG-IgG CBAs (median Spearman’s correlation coefficient R=0.866, range 0.806 to 0.961; figure 3 A) and a lower correlation between the fixed CBA and the live MOG-IgG CBAs (median R=0.800, range 0.778 to 0.809). There was no correlation between the MOG-IgG and MOG-IgM CBAs (median kappa value −0.071, range −0.134 to 0.179) or the MOG-IgG CBAs with ELISA (median R=0.094, range 0.060 to 0.273). These correlations are shown in more details in one illustrative assay per center (figure 3 B, assays Nr. 2, 4, 6, 7, 8). Although the separation of negative and positive samples was very good for the live CBAs (assays Nr. 2, 4, 6, and 7), the fixed assay (Nr. 8) was negative for 5 samples clearly positive in the live CBAs.

**Figure 3:**
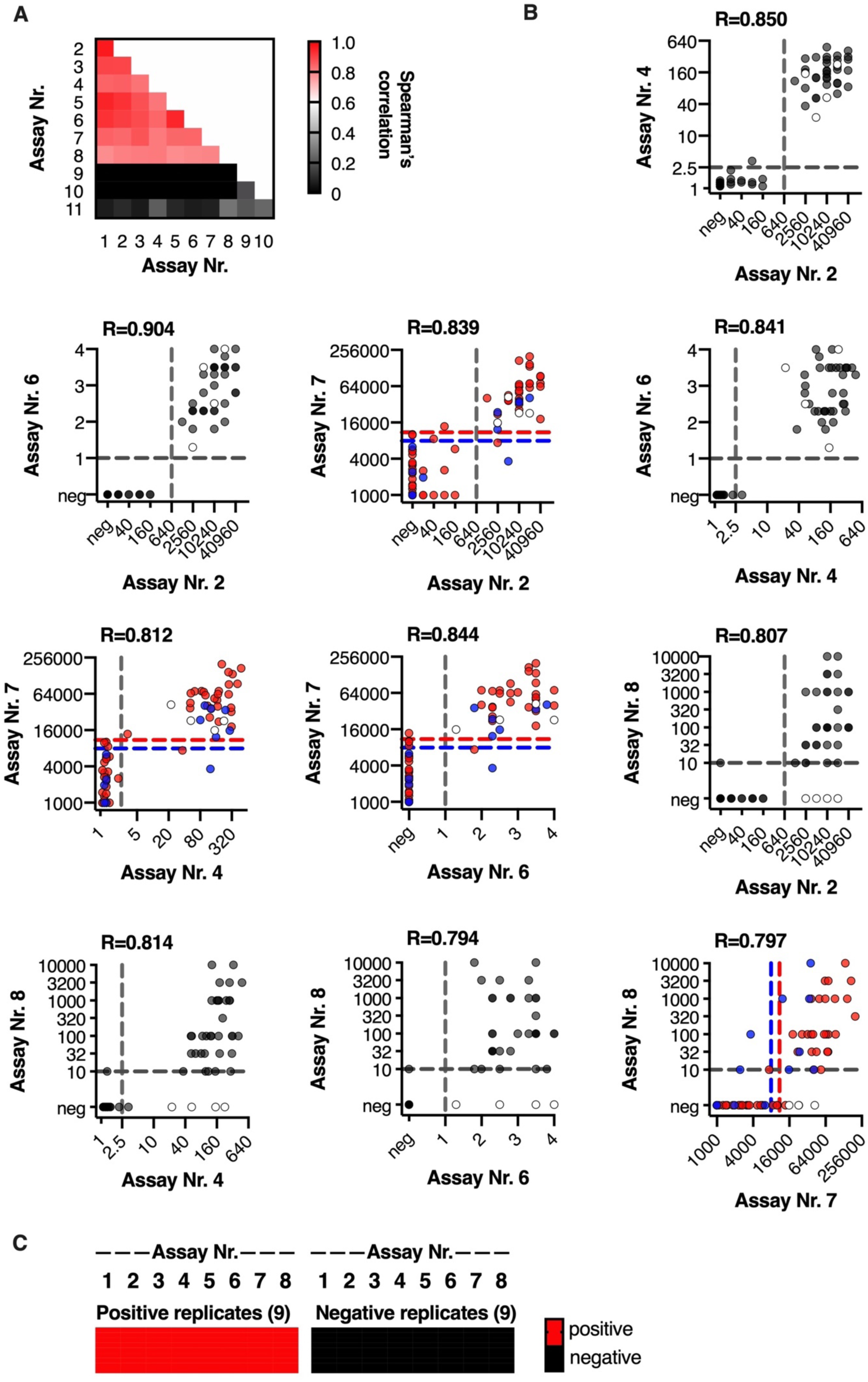
Correlation of quantitative values of clearly positive and negative samples and reproducibility of assay results. (A) A heatmap of Spearman’s correlation coefficients for all correlations. (B) Correlation of illustrative live (Nr. 2, 4, 6 and 7) and fixed CBAs (Nr. 8): Nr. 2 (center 1): CBA-IF IgG(Fc) titer (1:), Nr. 4 (center 2): CBA-FACS IgG1 binding ratio, Nr. 6 (center 3): CBA-IF IgG1 binding score, Nr. 7 (center 4): CBA-FACS IgG(H+L) delta mean fluorescence intensity, Nr, 8 (center 5): CBA-IF IgG(Fc) titer (1:). The cut-off values for all assays except assay Nr. 7 are indicated by the dashed grey lines. For assay Nr. 7 cut-off levels for pediatric samples (blue dots) are indicated by the blue dashed line and cut-off levels for adult samples (red dots) are indicated by the red dashed line. Samples which are positive in live CBAs but not the fixed CBA (assay Nr. 8) are indicated by the white dots. (C) Qualitative results for 9 positive and 9 negative samples from phase I re-tested in a blinded way by all assays in phase II.

As a technical control we included 10 samples containing humanized monoclonal MOG-IgG (5) or MOG-IgM (5) antibodies provided by center 5. Results are shown in supplementary results (efigure 2). Importantly, these humanized monoclonal antibodies were not recognized by some of secondary antibodies, particularly the anti-human IgG1 antibody. Moreover, the anti-human IgG(H+L), but not the IgG(Fc) secondary antibodies, also recognized the humanized monoclonal MOG-IgM at the lowest dilution as borderline negative for MOG-IgG.

### MOG-IgG results are reproducible within centers

All centers reproduced the MOG-IgG results from their samples submitted for phase I and the 9 positive and 9 negative replicates that were resent blinded and integrated into the cohort with the borderline samples in phase II (figure 3 C and supplementary results, efigure 3 and etable 3).

### CBAs for MOG-IgG show less agreement on low positive and borderline negative samples

In the second phase of this study (figure 1, phase II) we analyzed samples sent as low positive (n=39) or borderline negative (n=13) by the participating centers and 30 samples from healthy blood donors. Qualitative results obtained by the different CBAs for MOG-IgG are shown in figure 4 A and supplementary results, etable 3. In general, there was a good agreement for the eight MOG-IgG CBAs for the samples from blood donors: 29 of the 30 samples (97%) were negative in all eight CBAs and one sample was positive in four CBAs (figure 4 B).

**Figure 4:**
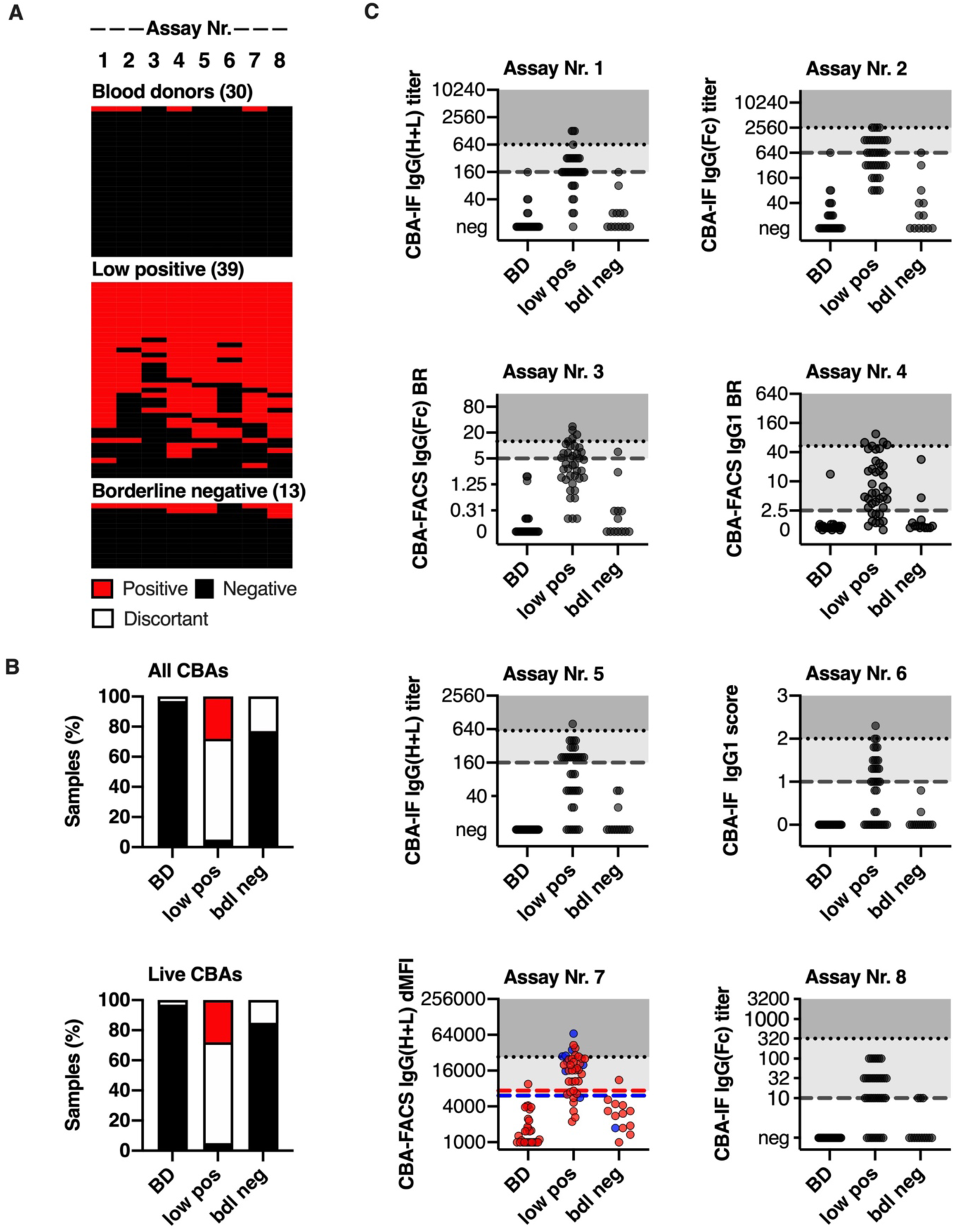
Qualitative and quantitative results of all MOG-IgG CBAs for blood donors (BD, n=30), low positive (n=39) and borderline negative (n=40) samples. (A) Qualitative results according to the serostatus sent. Each column is an individual MOG-IgG CBA and each row is an individual serum sample. Negative samples are black and positive samples are red. The samples are shown according to their serostatus sent by the individual centers. (B) Agreement of assay results for IgG CBAs (assays 1-8). Sample are grouped according to their agreement in all 8 CBAs or in the 7 live CBAs (red: positive in all CBAs, black: negative in all CBAs, white: discordant). (C) Quantitative results for the 8 MOG-IgG CBA-IF and CBA-FACS assays. The cut-off values for all assays except assay Nr. 7 are indicated by the dashed grey lines. For assay Nr. 7 cut-off levels for pediatric samples (blue dots) are indicated by the blue dashed line and cut-off levels for adult samples (red dots) are indicated by the red dashed line. The quantitative range of each assay result for its probability to be seropositive in all live CBAs is indicated by the dotted line and shaded in darker grey (100% probability), whereas the range of discordant samples is shaded in light grey. BD = blood donors, low pos = positive values just above the individual cut-off levels, bdl neg = borderline negative values just below the individual cut-off levels.

The agreement for low positive samples was less good: 2 of the 39 samples (5%) sent as borderline positive were negative in all 8 CBAs and only 11 samples (28%) were positive in all 8 CBAs. Therefore, the CBAs had a complete agreement of 33%. The remaining 26 samples (67%) were positive in 7 (n=8), 6 (n=2), 5 (n=6), 4 (n=2), 3 (n=3), 2 (n=2) and 1 (n=3) assays. The agreement for borderline negative samples was better: 10 of the 13 samples (77%) were negative in all 8 CBAs and no sample (0%) was positive in all 8 CBAs. Therefore, the CBAs had a complete agreement of 77%. The remaining three samples (23%) were positive in 7 (n=1), 3 (n=1), and 1 (n=1) assays.

The quantitative values for all MOG-IgG CBAs are provided in figure 4 C. From this figure it is evident that many of the positive signals are around the cut-off, and under the quantitative range described above for samples positive in all live CBAs.

Further, MOG-IgMs at a titer ≥ 1:160 were a rare finding in samples sent as low positive (n=5, all 1:160) and absent in samples sent as borderline negative or blood donors. These 5 samples were positive for MOG-IgG in 1 (n=1), 4 (n=1), 5 (=2) and 6 (n=1) CBAs.

### Samples identified as MOG-IgG positive in all CBAs are associated with a non-MS demyelinating disease course

None of the 13 clinically definite MS patients in the study or the five patients with other neurological diseases were within the 47 patients who tested positive on all live CBAs (supplementary results, efigure 4). These 47 patients had typical MOG-IgG associated clinical phenotypes such as optic neuritis, ADEM, myelitis, AQP4 seronegative NMOSD or other demyelinating phenotypes reported to be associated with MOG-IgG. The 32 discordant samples were from 27 patients with typical MOG associated clinical phenotypes mentioned above, but also from a healthy blood donor and four MS patients. Finally, the 82 samples negative in all live CBAs were from 24 patients with typical MOG associated clinical phenotypes, MS (9), other neurological diseases (10) and healthy controls (39).

## DISCUSSION

In this study we compared the reproducibility amongst the most frequently used assays for serum MOG-IgG detection, such as live and fixed CBA-IF, live CBA-FACS, and ELISA. Our data clearly indicate that strong positive and clearly negative samples are reproducible between centers where live cells expressing native full-length human MOG are used as the assay substrate. In the four different national testing centers using different live CBAs there was 96% concordance for all samples tested. The agreement was less good when a fixed CBA-IF (90%) tested in-house by the company (center 5) was included which is consistent with recent studies demonstrating that some conformational epitopes of MOG are lost upon fixation of MOG-expressing cells ^4, 17, 27^. Importantly, most of these discordant negative results on the fixed MOG-IgG assay had high MOG-IgG titers in live CBAs and were from typical MOG-IgG associated demyelinating syndromes. There is utility in the commercial fixed MOG-IgG testing in places where live MOG-IgG CBAs are unavailable, but this assay will miss 10-15% of positive cases. A recent study highlighted an issue with specificity in commercial MOG-IgG testing, particularly in samples that were only positive at low dilutions ^4^. Therefore, clinicians should consider retesting unexpected MOG-IgG results at centers offering live CBAs. Finally, ELISAs did not distinguish between the positive and negative patient samples and showed no concordance with CBAs for detection of human MOG-IgG conclusively demonstrating that ELISAs are not suitable for the detection of human MOG-IgG. Although this has been shown in several studies (summarized in ^29^), some laboratories still use this method for MOG-IgG detection. We hope that our findings inform neurologists that only CBAs should be used for the measurement of human MOG-IgG. Moreover, and in agreement with previous studies ^4, 17, 27^, live CBAs remain the gold standard for the detection of human MOG-IgG.

The agreement of MOG-IgG CBAs for low positive sample was less good (33% concordance) and MOG-IgG assays were particularly discordant at the borderline of positivity. This raises the pertinent question where to draw cut-off values and how they influence the clinical interpretation of diagnostic results. If we examine the clinical phenotype of people with high MOG-IgG levels, which are consistently detected by all CBAs, we identify patients with non-MS demyelinating phenotypes (such as ADEM, NMOSD, optic neuritis, myelitis, and other demyelinating diseases) ^1^. In contrast, the low positive samples which showed a lack of reproducibility between centers had a wider range of clinical phenotypes that mostly include the same phenotypes (ADEM, NMOSD, optic neuritis, myelitis and other demyelinating diseases), but also a proportion of every control group (clinically definite MS, other neurological diseases and healthy individuals) making their interpretation difficult. It is unlikely that lower levels of pathogenic antibody cause a wider disease presentation suggesting that some of these phenotypes are not driven by MOG-IgG. Hence an argument can be made that the presence of low positive MOG-IgG is only meaningful in the “correct clinical context” such as in patients with ON, myelitis, ADEM or encephalitis but not in the context of other diseases, particularly MS,^1, 20, 32^. This is a circular but reasonable interpretation of low positive results, but with caveats. There will be a false-positive rate even within the correct clinical context that should be considered and an estimate of this would be useful for any test. Secondly, clinical criteria are not perfect. There are individuals who fulfill criteria for MS, but are often atypical; perhaps the MOG-IgG result has utility in this context in ruling out MS, and should not be ignored out of hand. Importantly, when extrapolating from experiences on the treatment of NMOSD and a recent larger study on treatment of patients with MOG-IgG from France, disease modifying treatments for MS may not work in MOG-IgG positive patients and may even exacerbate disease ^1, 32-34^. The third interpretation is that these low positive results that are not reproducible between centers are not useful clinically and in fact dilute the utility of a more specific test. Finally, as a general consideration in samples not taken at disease onset, other confounding factors such as preceding steroid use or other immunosuppressive treatments and remission could lower a positive MOG-IgG result. It is important to note that MOG-IgG levels are often non-normally distributed in large patient cohorts, and a skewing towards these lower MOG-IgG titers has been observed in many studies ^1^.

Further work is now needed to better define the most useful clinical cut-off and to establish if there is any added benefit in identifying patients with “low positive” MOG-IgG. We propose that this should be done in a collaborative effort. We need to better characterize “false positive” cases, such as classical MS cases, other neurological diseases, and healthy individuals, and get more information on the clinical sensitivity and specificity of all assays by using appropriate controls, such as systemic autoimmune diseases, non-inflammatory neurological controls, and healthy controls. It is of great interest to establish how these antibodies relate to different clinical phenotypes and whether they are a mixture of pathogenic and bystander antibodies that all bind MOG in vitro.

To conclude we have shown that currently used live CBAs to measure MOG-Abs showed excellent agreement for clearly positive and negative samples, but low positive samples were more discordant. Further work is now required to standardize the clinically most useful assay.

## Abbreviations

ADEM: acute disseminated encephalomyelitis
AQP4: aquaporin-4
CBA: cell-based assay
ELISA: enzyme linked immunosorbent assay
FACS: fluorescence activated cell sorting
IF: immunofluorescence
MOG: myelin oligodendrocyte glycoprotein
MS: multiple sclerosis
NMOSD: neuromyelitis optica spectrum disorder
ON: optic neuritis.

## APPENDIX AUTHORS

| Name | Location | Role | Contribution |
| --- | --- | --- | --- |
| Markus Reindl | Medical University of Innsbruck, Austria | Author | Designed and conceptualized the study; analyzed and interpreted the data; and drafted and revised the manuscript for intellectual content |
| Kathrin Schanda | Medical University of Innsbruck, Austria | Author | Analyzed the data; revised the manuscript for intellectual content |
| Mark Woodhall | University of Oxford, Oxford, UK | Author | Analyzed the data; revised the manuscript for intellectual content |
| Fiona Tea | University of Sydney, Sydney, Australia | Author | Analyzed the data; revised the manuscript for intellectual content |
| Sudarshini Ramanathan | University of Sydney, Sydney, Australia | Author | Analyzed the data; revised the manuscript for intellectual content |
| Jessica Sagen | Mayo Clinic, USA | Author | Analyzed the data; revised the manuscript for intellectual content |
| Jim Fryer | Mayo Clinic, USA | Author | Analyzed the data; revised the manuscript for intellectual content |
| John Mills | Mayo Clinic, USA | Author | Analyzed the data; revised the manuscript for intellectual content |
| Bianca Teegen | Euroimmun AG, Germany | Author | Analyzed the data; revised the manuscript for intellectual content |
| Swantje Mindorf | Euroimmun AG, Germany | Author | Analyzed the data; revised the manuscript for intellectual content |
| Nora Ritter | Euroimmun AG,<br>Germany | Author | Analyzed the data;<br>revised the<br>manuscript for<br>intellectual content |
| Ulrike Krummrei | Euroimmun AG,<br>Germany | Author | Analyzed the data;<br>revised the<br>manuscript for<br>intellectual content |
| Winfried Stöcker | Euroimmun AG,<br>Germany | Author | Analyzed the data;<br>revised the<br>manuscript for<br>intellectual content |
| Juliane Eggert | Institute for Quality<br>Assurance,<br>Germany | Author | Analyzed the data;<br>revised the<br>manuscript for<br>intellectual content |
| Eoin Flanagan | Mayo Clinic, USA | Author | Provided reagents<br>and data; revised<br>the manuscript for<br>intellectual content |
| Melanie Ramberger | Medical University of<br>Innsbruck, Austria<br>and University of<br>Oxford, Oxford, UK | Author | Provided reagents<br>and data; revised<br>the manuscript for<br>intellectual content |
| Harald Hegen | Medical University of<br>Innsbruck, Austria | Authors | Provided reagents<br>and data; revised<br>the manuscript for<br>intellectual content |
| Kevin Rostasy | Witten/Herdecke<br>University, Germany | Author | Provided reagents<br>and data; revised<br>the manuscript for<br>intellectual content |
| Thomas Berger | Medical University of<br>Vienna, Austria | Author | Provided reagents<br>and data; revised<br>the manuscript for<br>intellectual content |
| M. Isabel Leite | University of Oxford,<br>Oxford, UK | Author | Provided reagents and data; revised the manuscript for intellectual content |
| Jacqueline Palace | University of Oxford,<br>Oxford, UK | Author | Provided reagents and data; revised the manuscript for intellectual content |
| Sarosh R Irani | University of Oxford,<br>Oxford, UK | Author | Provided reagents and data; revised the manuscript for intellectual content |
| Russell Dale | University of<br>Sydney, Sydney,<br>Australia | Author | Provided reagents and data; revised the manuscript for intellectual content |
| Monika Probst | Institute for Quality<br>Assurance,<br>Germany | Author | Designed and conceptualized the study; analyzed and interpreted the data; revised the manuscript for intellectual content |
| Christian Probst | Euroimmun,<br>Germany | Author | Analyzed the data; revised the manuscript for intellectual content |
| Fabienne Brilot | University of<br>Sydney, Sydney,<br>Australia | Author | Designed and conceptualized the study; analyzed and interpreted the data; and drafted and revised the manuscript for intellectual content |
| Sean Pittock | Mayo Clinic, USA | Author | Designed and conceptualized the study; analyzed and interpreted the data; and drafted and revised the manuscript for intellectual content |
| Patrick Waters | University of Oxford, Oxford, UK | Author | Designed and conceptualized the study; analyzed and interpreted the data; and drafted and revised the manuscript for intellectual content |

## SUPPLEMENTARY INFORMATION

### 1. Supplementary methods for Center 1 (Medical University of Innsbruck, Austria) *Assays Nr. 1, 2 and 9 (live CBA-IF)*

For this study we used our live CBA-IF MOG-Ab assay as initially described in 2011 ^2, 5^ and now widely used not only by our group, but also by other laboratories (***assay Nr. 1***). Our MOG-Ab assay was reevaluated in a blinded fashion by the German NEMOS study group in 2016 ^14^, with an excellent specificity of 99.5% (95% CI 0.97 to 1.00) in 222 controls (MS, neurological and healthy controls). Further, a large blinded study using our MOG-Ab assay revealed a specificity of 100% in 200 patients with chronic progressive MS ^35^.

In response to a report of possible co-detection of IgM MOG-Ab with the anti-IgG (H+L) secondary antibody used in our original assay ^3^, we have refined our assay by using a human IgG(Fc)-specific secondary antibody instead of an anti-human IgG(H+L) secondary antibody^14, 15^.

For all CBAs HEK-293 cells were grown in 75cm^2^ flasks using DMEM culture medium containing 4.5g/l glucose, 10% FCS, 4mM L-glutamine (Life Technologies) and 1xnon-essential amino acids (NEAA, Life Technologies) and passaged every 3 days.

HEK-293 cells were seeded into 96-well cell culture plates (TPP) at a density of 200.000 cells/ml using the culture medium mentioned above. 24h after seeding, transfection was performed using the pEGFP-N1-hMOGalpha1 plasmid and the transfection reagent Fugene HD (Promega). Cells were maintained in a humified incubator until commencing the assay.

In principal, the set-up of the different analysis was designed to avoid multiple thawing cycles: Therefore, sera were first screened at a dilution of 1:20 and 1:40 using the anti-human IgG H+L antibody (*assay Nr. 1*) and in parallel in serial dilution 1:20 to 1:160 using the human anti-MOG IgMµ-specific antibody (*assay Nr. 9*) as most patient IgM titers can be found in this range according to our experience. After detection of a positive signal during the screening procedure, sera were retested using serial dilutions in two-fold steps starting at 1:20. Human anti-MOG IgG Fc-specific testing (*assay Nr. 2*) was performed according to the screening result of the IgG H+L result: If negative, the negative result was confirmed by sera dilutions 1:20 and 1:40. In case of positive IgG H+L signals, serial titrations were carried out in parallel to the IgG H+L titrations and (possible) IgMµ titrations to thaw the sample only 2 times for all CBA-IF analysis. Briefly, 24h post-transfection, plates were removed from the incubator and the culture medium was replaced by blocking solution, consisting of 0.2 µg/ml goat IgG (Sigma) in 10% heat-inactivated FCS in PBS (Sigma, assay buffer). After 10 minutes, blocking was removed and diluted samples were added, followed by an incubation for 1h at 4°C. Thereafter, cells were washed three times using assay buffer, followed by incubation with the appropriate secondary antibody (Cy 3TM-conjugated anti-human IgG H+L, Jackson ImmunoResearch 109-166-088, 1:200; Alexa Fluor®594-conjugated goat anti-human IgG Fcγ Fragment specific, Jackson ImmunoResearch 109-586-098, 1:750; Alexa Fluor®594-conjugated goat anti-human IgM Fc_5µ_ Fragment specific, Jackson ImmunoResearch 109-585-129, 1:750; all antibodies were diluted in assay buffer) for 30 minutes at room temperature. Cells were washed three times with assay buffer and finally DAPI (0.1 µg/ml in assay buffer, Sigma) was added to indicate dead cells. Screening and determination of titer levels was performed by two investigators, both using individual result sheets and being blinded to the results of the other. The microscope was a Leica 4000B with a BGR filter. End-point titer levels were defined by the last dilution at which a specific signal was observed. Concordance rate between raters was 100%.

Assays Nr. 2, 3 and 9 were validated using 322 serum samples from people with inflammatory demyelinating diseases and healthy controls. eFigure 1 and eTable 1 show the results for the initial validation of this assay including the definition of the cut-off value and the comparison with our original IgG(H+L) assay. From eTable 1 it is evident that the specificity of assay Nr. 2 is higher than that of our initial MOG-Ab assay (assay Nr. 1), which is mainly due to the elimination of cross-reactive IgM seropositive samples.

**eTable 1.** Characterization of assays Nr. 1, 2, 3 and 9 for serum MOG-Ab.

|  | **CBA-IF**<br>**IgG(H+L)**<br>**Cut-off value**<br>**≥ 1:160**<br>**Assay Nr .1** | **CBA-IF**<br>**IgG(Fc)**<br>**Cut-off value**<br>**≥ 1:640**<br>**Assay Nr. 2** | **CBA-FACS**<br>**IgG(Fc)**<br>**Cut-off value**<br>**≥ 5**<br>**Assay Nr. 3** | **CBA-IF**<br>**IgM**<br>**Cut-off value**<br>**≥ 1:160**<br>**Assay Nr .9** |
| --- | --- | --- | --- | --- |
| **Optic neuritis** | 32 / 35 (91%) | 30 / 35 (86%) | 28 / 35 (80%) | 2 / 35 (6%) |
| **Myelitis** | 10 / 11 (91%) | 9 / 11 (82%) | 7 / 11 (64%) | 1 / 11 (9%) |
| **ADEM / MDEM** | 46 / 57 (81%) | 45 / 57 (78%) | 41 / 57 (72%) | 9 / 57 (16%) |
| **AQP4-Ab**<br>**negative NMOSD** | 19 / 22 (86%) | 17 / 22 (77%) | 12 / 22 (55%) | 2 / 22 (9%) |
| **AQP4-Ab positive**<br>**NMOSD** | 0 / 17 (0%) | 0 / 17 (0%) | 0 / 17 (0%) | 1 / 17 (6%) |
| **Multiple sclerosis** | 3 / 120 (2%) | 0 / 120 (0%) | 2 / 120 (2%) | 6 / 120 (5%) |
| **Healthy controls** | 1 / 60 (2%) | 0 / 60 (0%) | 2 / 60 (3%) | 7 / 60 (12%) |

ADEM = acute disseminated encephalomyelitis, MDEM = multiphasic disseminated encephalomyelitis, AQP4 = aquaporin 4, NMOSD = neuromyelitis spectrum disorder.

**eFigure 1:**
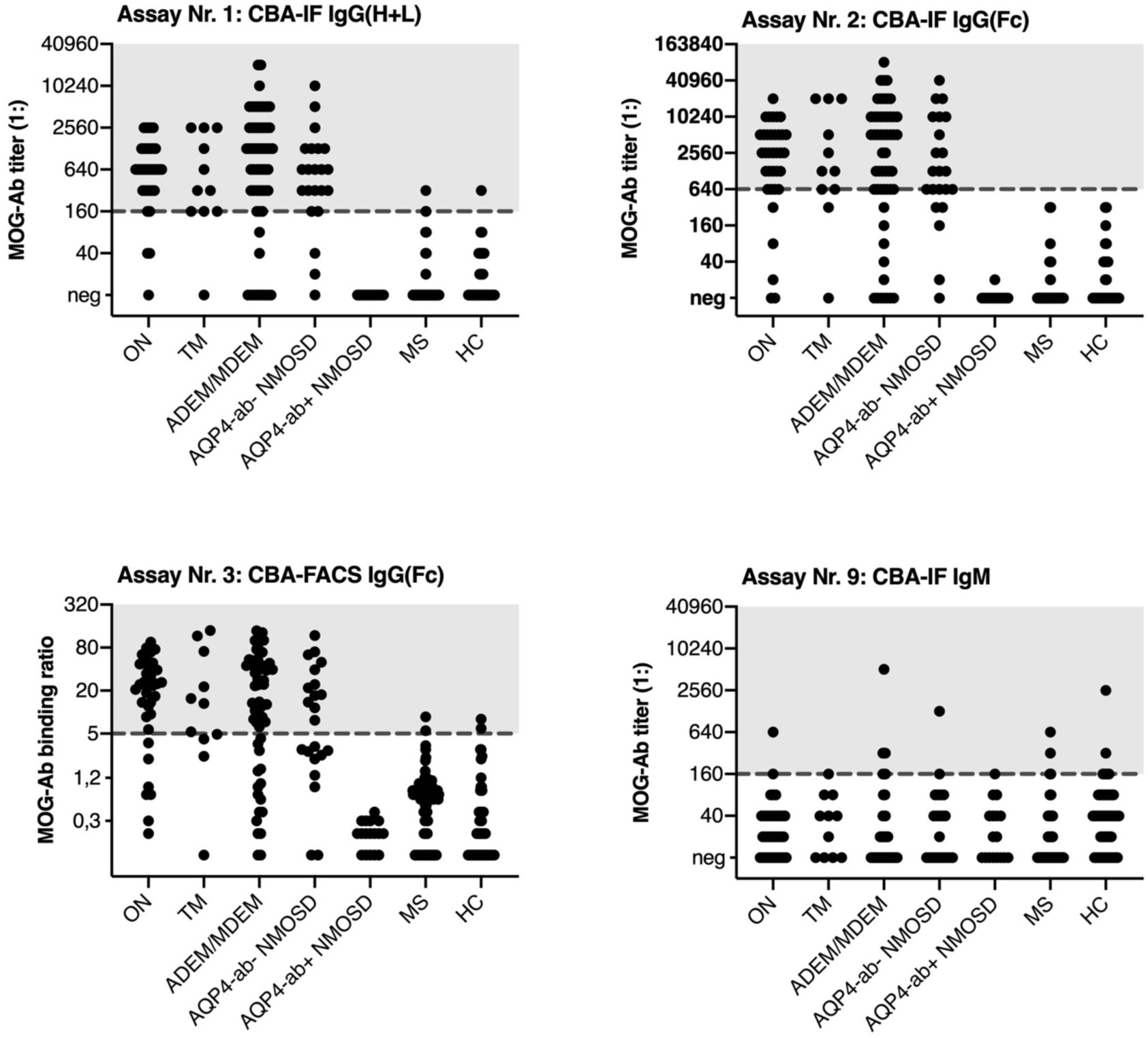
Characterization and validation of assays Nr. 2, 3 and 9 and comparison with assay Nr. 1 for serum MOG-Ab. Quantitative results for the all assays according to clinical diagnosis. Cut-off values are indicated by the dashed grey lines and positive samples are highlighted in grey.

### Assay Nr. 3 (live CBA-FACS)

HEK293 cells were transduced with commercially purchased adeno-associated virus (AAV) vectors containing AAV2-CMV-hMOG-GFP (SignaGen Laboratories, Rockville, MD, USA). According to the maufacturer’s guidelines, 0.5 Mio cells were seeded in a 6-well plate and infected with virus at a concentration of 0.1 Mio multiplicity of infection (MOI), stored and incubated overnight at 37°C and 8% CO2. Cells were then transferred into new culture medium and transduction efficacy was measured by fluorescent activated cell sorting measuring the expressed GFP signal. Transduced cells were cultured for two weeks and subsequently single-cell sorted by our core facility. GFP expressing single cells were collected in a 96-well plate and successively expanded. All analysis was performed with a monoclonal cell line derived from one single AAV-MOG transduced clone showing surface expression of MOG and good GFP expression.

The CBA-FACS assay was performed using HEK293 (HEK) cells and AAV-MOG-GFP transduced HEK293 (AAV-MOG) cells. For the assay, both cell lines were trypsinized, counted and mixed equally (1+1) to a density of 2 Mio/ml in 10% heat-inactivated FCS in PBS containing 1mM EDTA (FCS-EDTA). The cell mixture was placed on an upside-down rotator in the dark at room temperature for one hour of recovery. Sera were spun down at 10000g for 5min and diluted 1:50 in FCS-EDTA buffer. After recovery, 100µl/well of cells were transferred into a round-bottom 96 well plate (TPP) and 100µl/well of sera added in duplicates, giving a final serum dilution of 1:100. In addition to the analyzed samples, three control sera (high, medium, negative), as well as a calibrator serum and blank were added to each plate. The calibrator was placed in two duplicates on different positions on the plate to control for and ensure equal conditions for all samples. After one hour at 4°C on a horizontal shaker, cells were washed three times with FCS-EDTA and the secondary anti-human IgG Fc-specific APC-labelled antibody (1:200, Jackson ImmunoResearch 109-135-098) was added (100µl/well) for 30min at room temperature in the dark on a horizontal shaker. After two more washing steps, the cell pellet was dissolved in 30µl/well 1mM EDTA in PBS and 100µl/well cell-fix solution (BD) were added. Cells were fixed at 4°C for 15min. Measurement was performed on an Accuri C6 flow cytometer (BS), gating on the cell population as a whole and further gating on HEK only and AAV-MOG populations, restricting uptake to 10000 AAV-MOG cells. For calculations, the FL4 median fluorescence intensity (MFI) was used for both selected populations. The calculation for the final values was performed by first calculating the mean of the sample duplicate, followed by subtracting the MFI (HEK) of the MFI (AAV-MOG), resulting in the delta MFI of the sample. Finally, each sample delta MFI was calibrated with the delta MFI of the calibrator, therefore compensating for possible differences in test performance per day. Controls and the second calibrator were used as additional quality controls of each assay. Over the course of three years of performing the FACS assay the following control sample results were observed: negative control calibrated delta MFI mean 0.16 (standard deviation 0.04; coefficient of variation=23.04%), positive control calibrated delta MFI mean 19.8 (standard deviation 3.17; coefficient of variation=15.98%) and strong positive control calibrated delta MFI mean 54.4 (standard deviation 8.94; coefficient of variation=16.43%).

Assay Nr. 3 was validated using the same 322 serum samples from people with inflammatory demyelinating diseases and healthy controls as assay Nr. 2. eFigure 1 and eTable 1 show the results for the initial validation of this assay including the definition of the cut-off value and the comparison with our original IgG(H+L) assay. From this data we determined a cut-off level of ≥5.0 calibrated delta MFI using ROC analysis.

### Assay Nr. 10 (ELISA)

ELISA was performed using the commercially available ANASPEC SensoLyteR anti-Human MOG (1-125) Specific Quantitative ELISA Kit (lots 1012 and 1013, by ANASPEC EGT group) according to the manufacturer’s instruction. All samples were analyzed in duplicates. Sera were used at a dilution of 1:40 and the anti-human IgG-HRP (Component G) secondary antibody was used at a dilution of 1:2000 for 45min. Absorbance (OD) was measured at 450nm. Calculation of ng/ml was performed using a polynomal calibration curve (standard curve: R^2^=0.99 in all plates assayed). Samples were determined to be positive at ≥150ng/ml as described before ^30^.

### 2. Supplementary results

As a technical control we have also included 10 samples containing monoclonal humanized IgG or IgM antibodies provided by center 5 in phase 1. The humanized monoclonal MOG-Ab 8-18-C5 ^31^ (expressed as human IgG1 or IgM) was used in increasing dilutions to assess the technical sensitivity of assays. This antibody was not recognized in assays 4, 5, 6, 9 or 11. Detailed results are shown in efigure 2.

However, these results must be interpreted with caution since the humanized IgG1 MOG-Ab was not recognized by some of the secondary antibodies, particularly the anti-IgG1 antibodies. Furthermore, only dilutions, but no concentration, of the monoclonal antibodies were given and the final dilutions varied greatly between assays.

All centers reproduced the MOG-IgG results from their samples submitted for phase I (efigure 3).

**eFigure 2:**
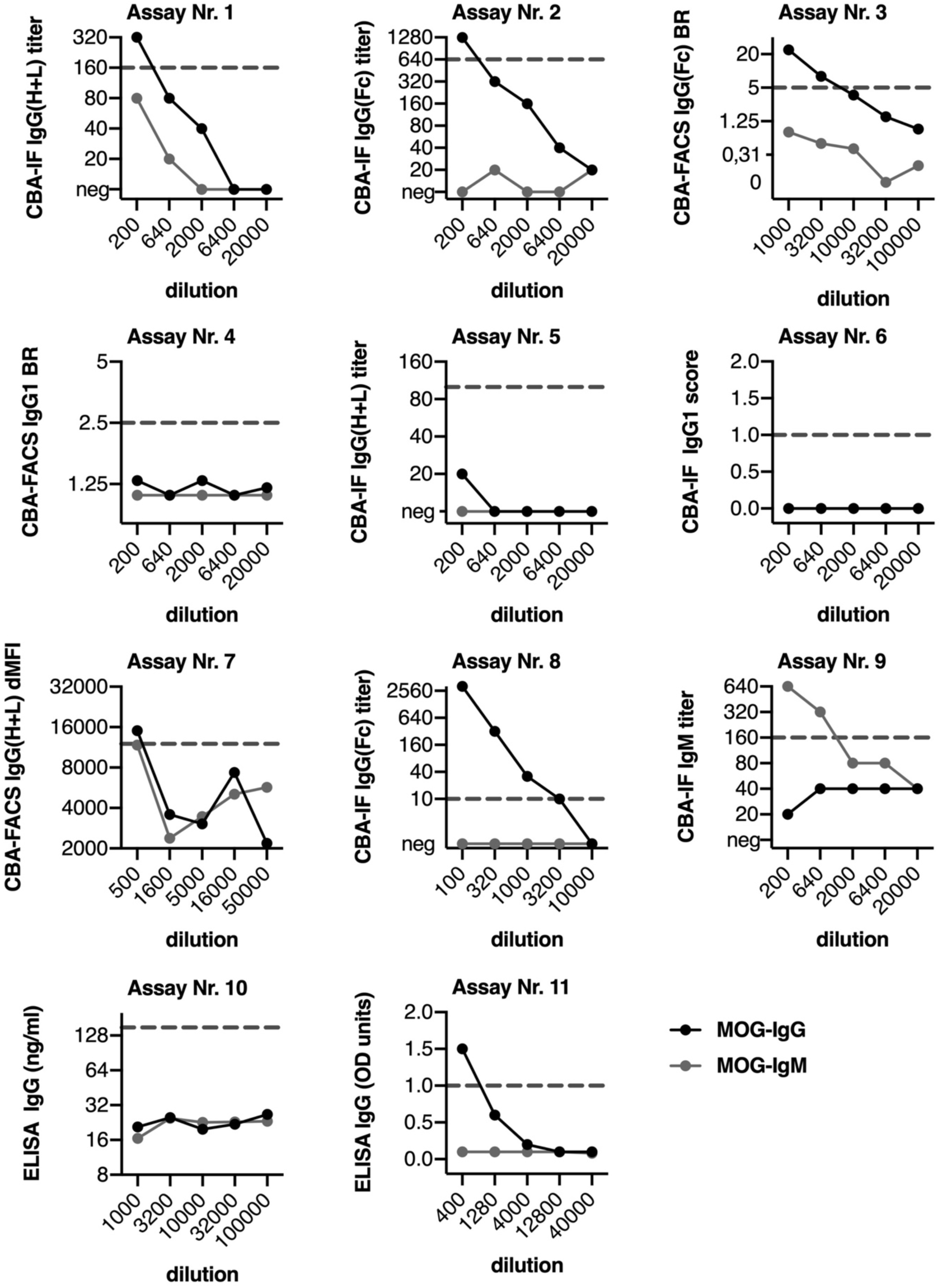
Quantitative results for all assays for monoclonal humanized IgG or IgM antibodies according to the endpoint dilution of samples for each assay. The cut-off values for all assays are indicated by the dashed grey lines.

**eFigure 3:**
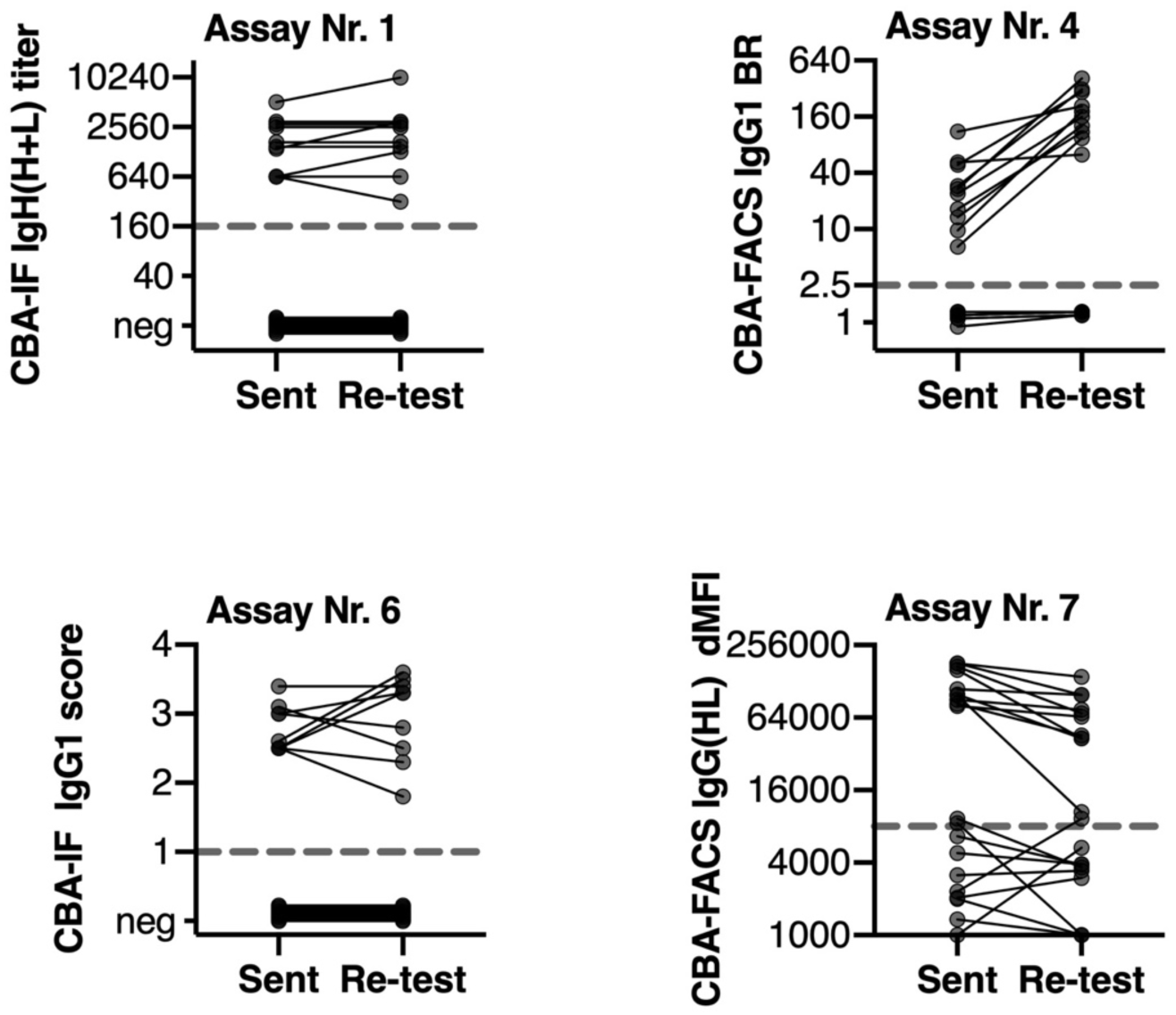
Reproducibility of CBAs for MOG-IgG. (A) Quantitative results according to the quantitative values sent and re-tested by assay Nr. 1, Nr. 4, Nr. 6 and Nr. 7. BR=binding ratio, dMFI = delta mean fluorescence intensity.

**eFigure 4:**
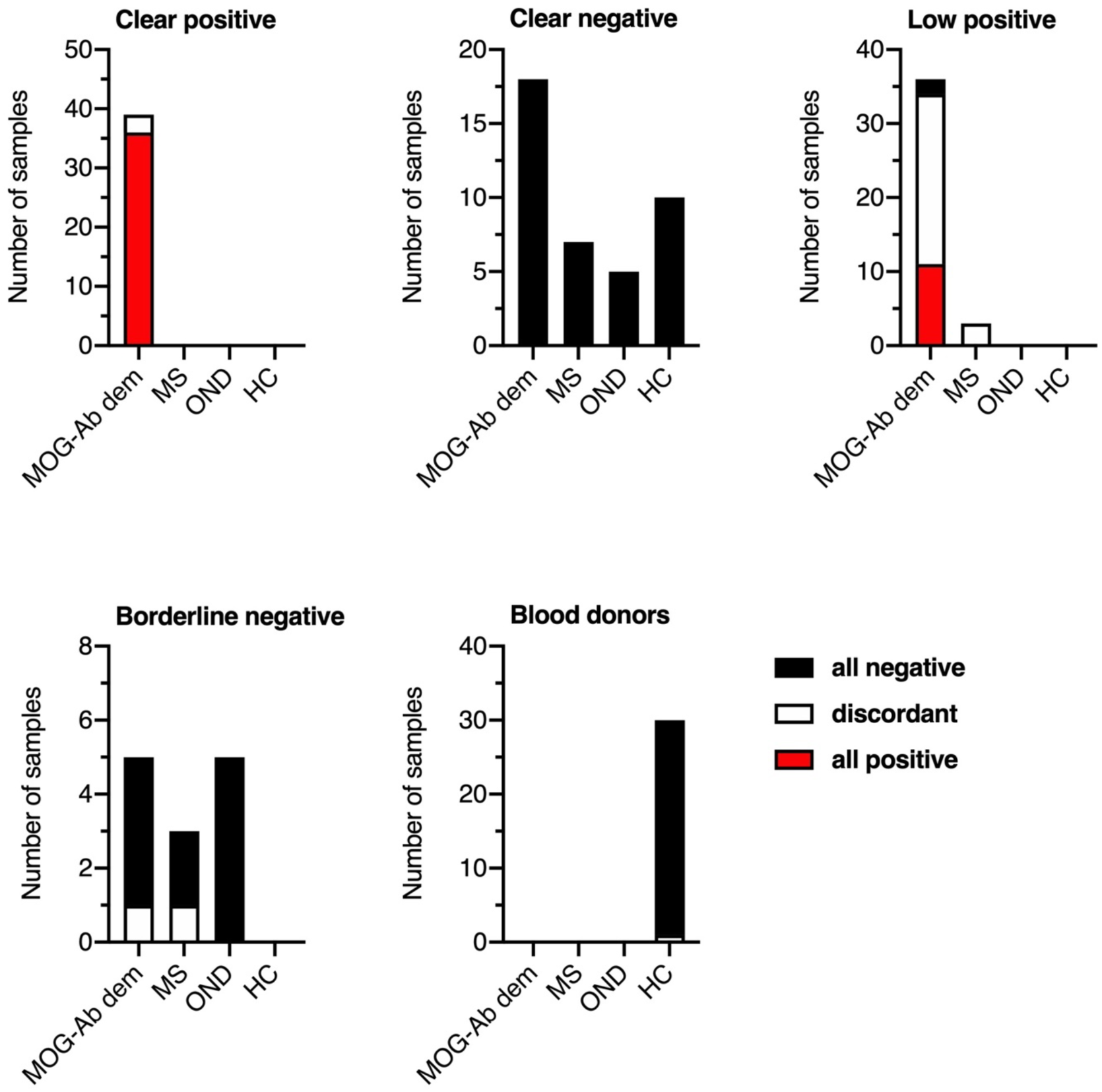
Clinical phenotypes of samples according to the agreement of results in the 7 live CBAs for MOG-IgG antibodies and serostatus sent. Results (in number of all samples) are grouped according to their agreement in all 7 live CBAs (red: positive in all live CBAs, black: negative in all live CBAs, white: discordant). MOG-Ab dem = typical MOG-IgG associated clinical phenotypes such as optic neuritis, ADEM, myelitis, AQP4 seronegative NMOSD or other demyelinating phenotypes reported to be associated with MOG-IgG; MS = multiple sclerosis; OND = other neurological diseases; HC = healthy controls.

**eTable 2:** Serostatus (quanititative range) of samples sent by centers 1-4.

|  | Center 1 (Innsbruck, assay Nr. 1) | Center 2 (Mayo Clinic, assay Nr. 4) | Center 3 (Oxford, assay Nr. 6) | Center 4 (Sydney, assay Nr. 7) |
| --- | --- | --- | --- | --- |
| Cut-off value | $\geq 1:160$ | $\geq 2.5$ | $\geq 1$ | Dependent on day of assay, determined on 24-aged matched controls |
| Sent as clear positive | N=10<br>1280 (640-5120) | N=10<br>25.4 (6.5-111.0) | N=9<br>2.5 (2.2-3.5) | N=10<br>104000 (83000-180000) |
| Sent as clear negative | N=10<br>0 (0) | N=10<br>1.2 (0.9-1.3) | N=10<br>0 (0) | N=10<br>2730 (60-9300) |
| Sent as low positive | N=10<br>160 (160-320) | N=11<br>3.1 (2.8-5.9) | N=8<br>1 (1-1.5) | N=10<br>26491 (15840-56738) |
| Sent as borderline negative | N=0 | N=0 | N=3<br>0 (0) | N=10<br>3524 (567-6530) |

Data are shown as median (range). Assay Nr. 1 live CBA-IF IgG(H+L), titer (1:); assay Nr. 4 live CBA-FACS IgG1, FACS binding ratio; assay Nr. 6 live CBA-IF IgG1, binding score; Assay Nr. 7 live CBA-FACS IgG(H+L), delta mean fluorescence intensity.

**eTable 3:**
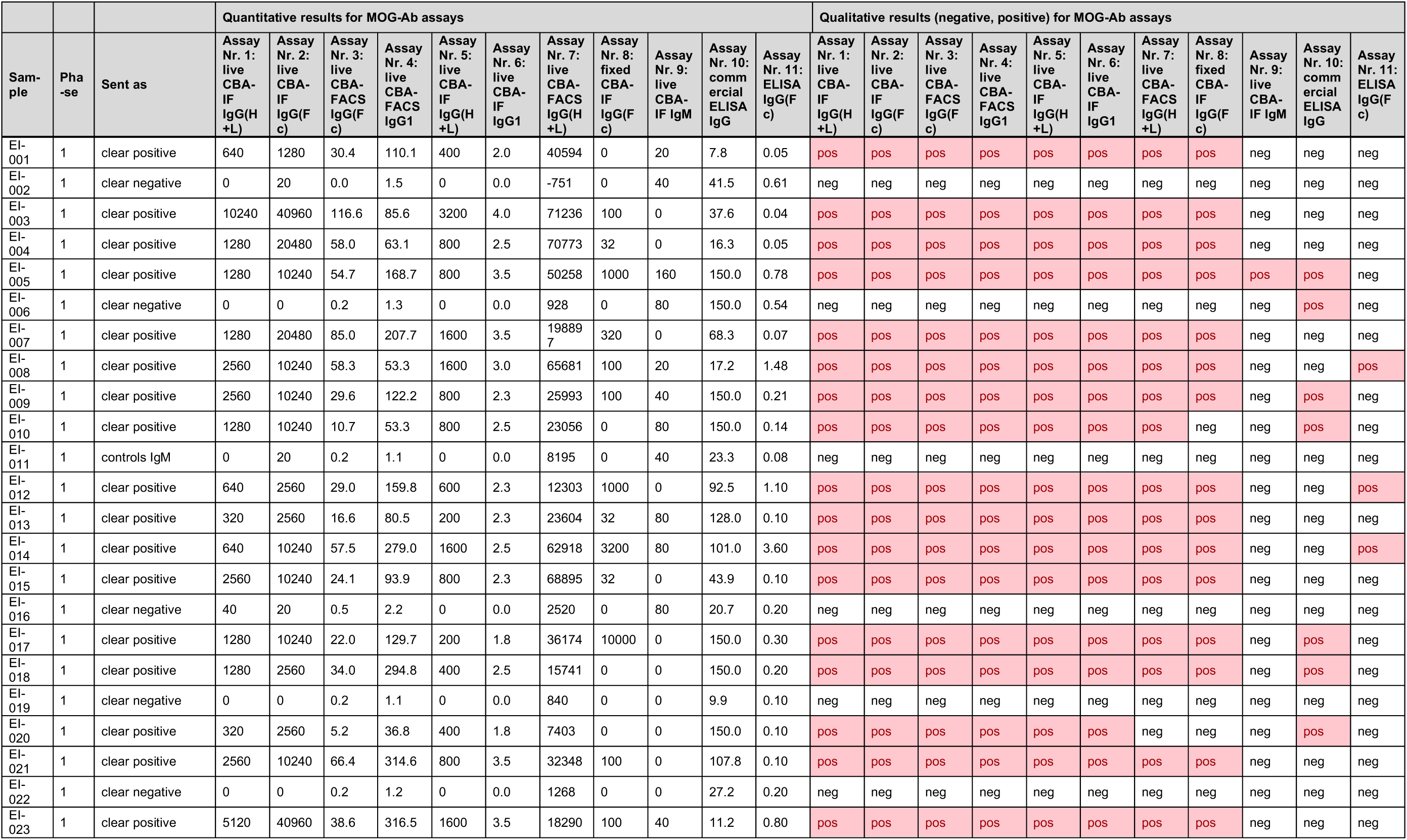
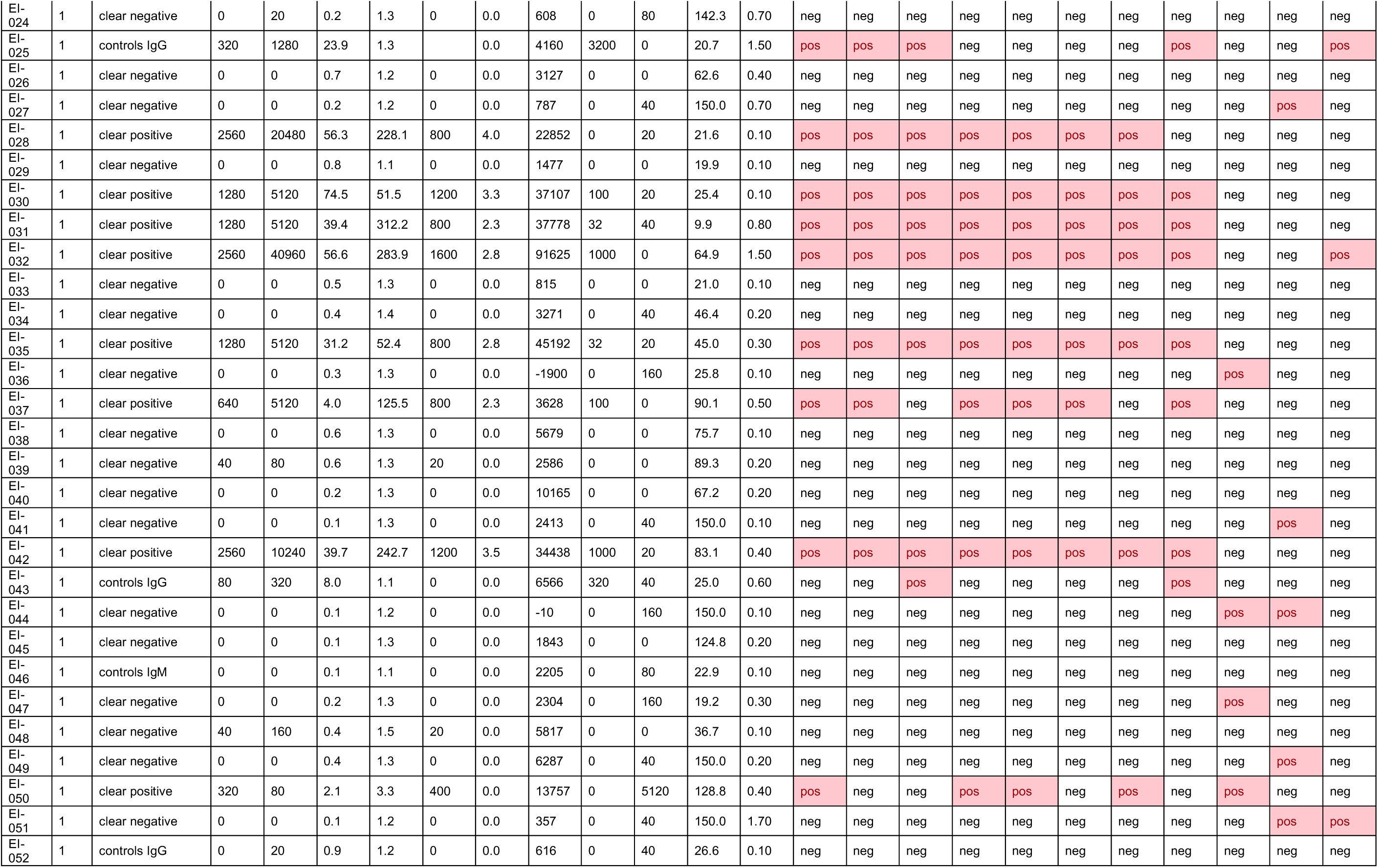
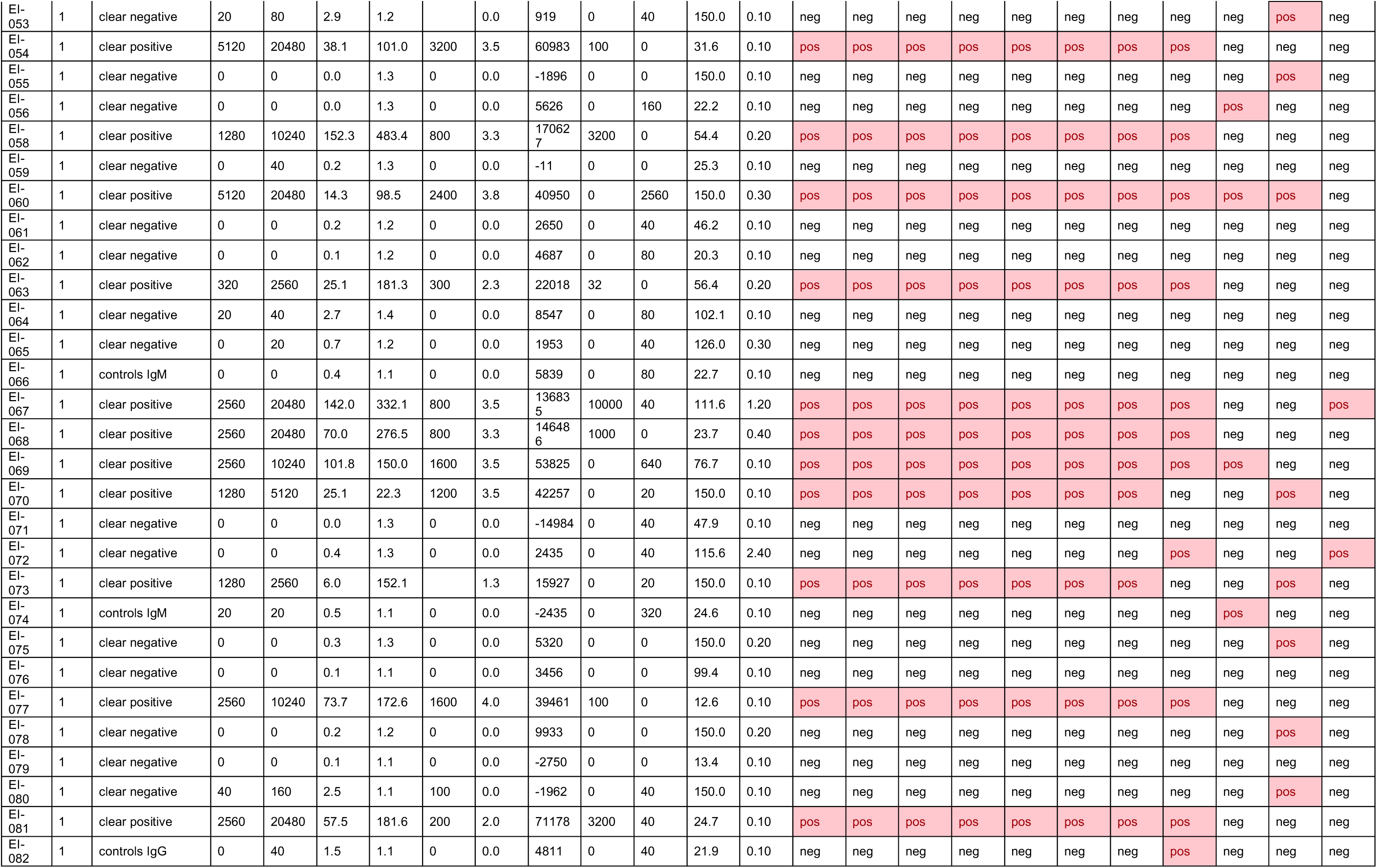
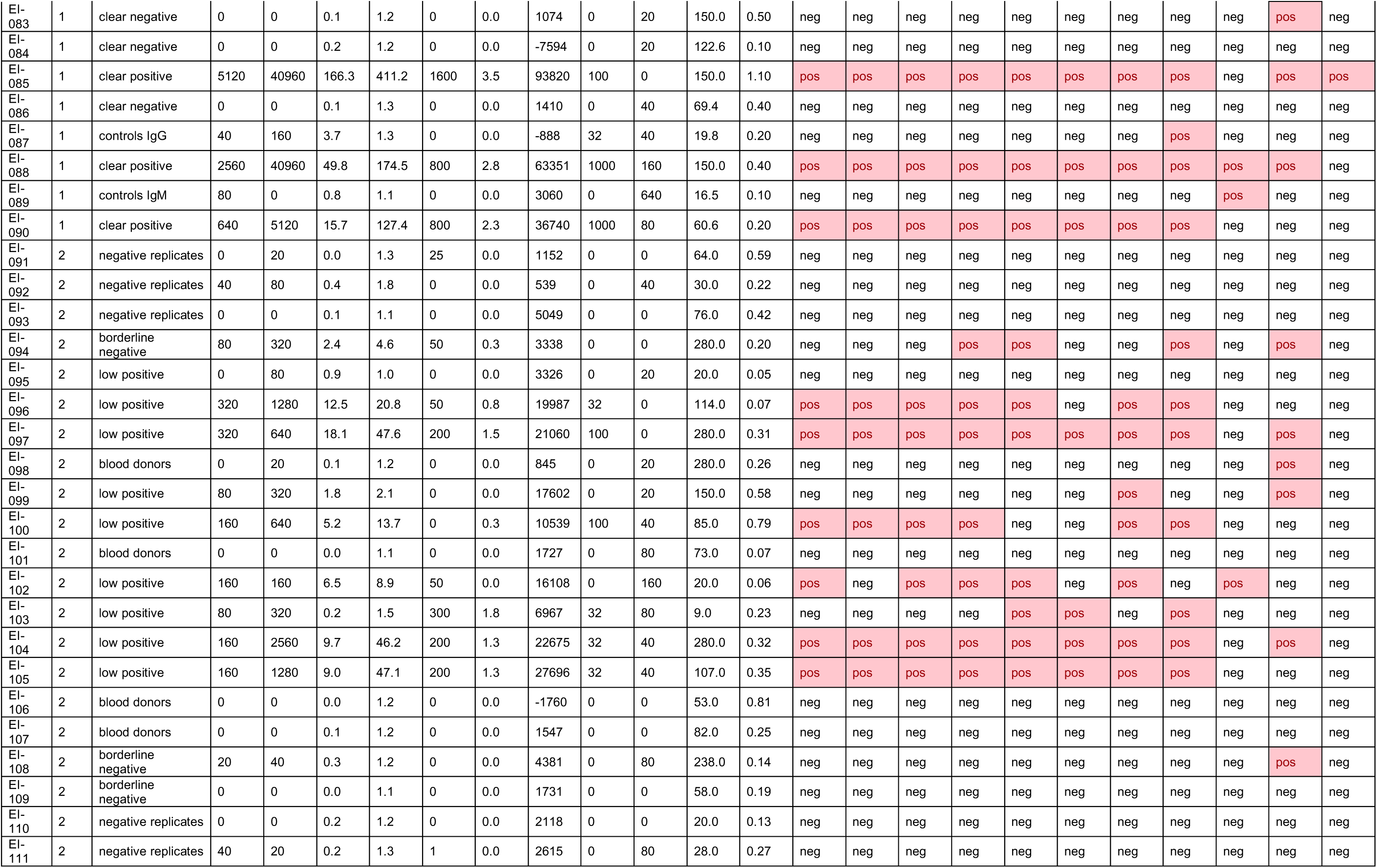
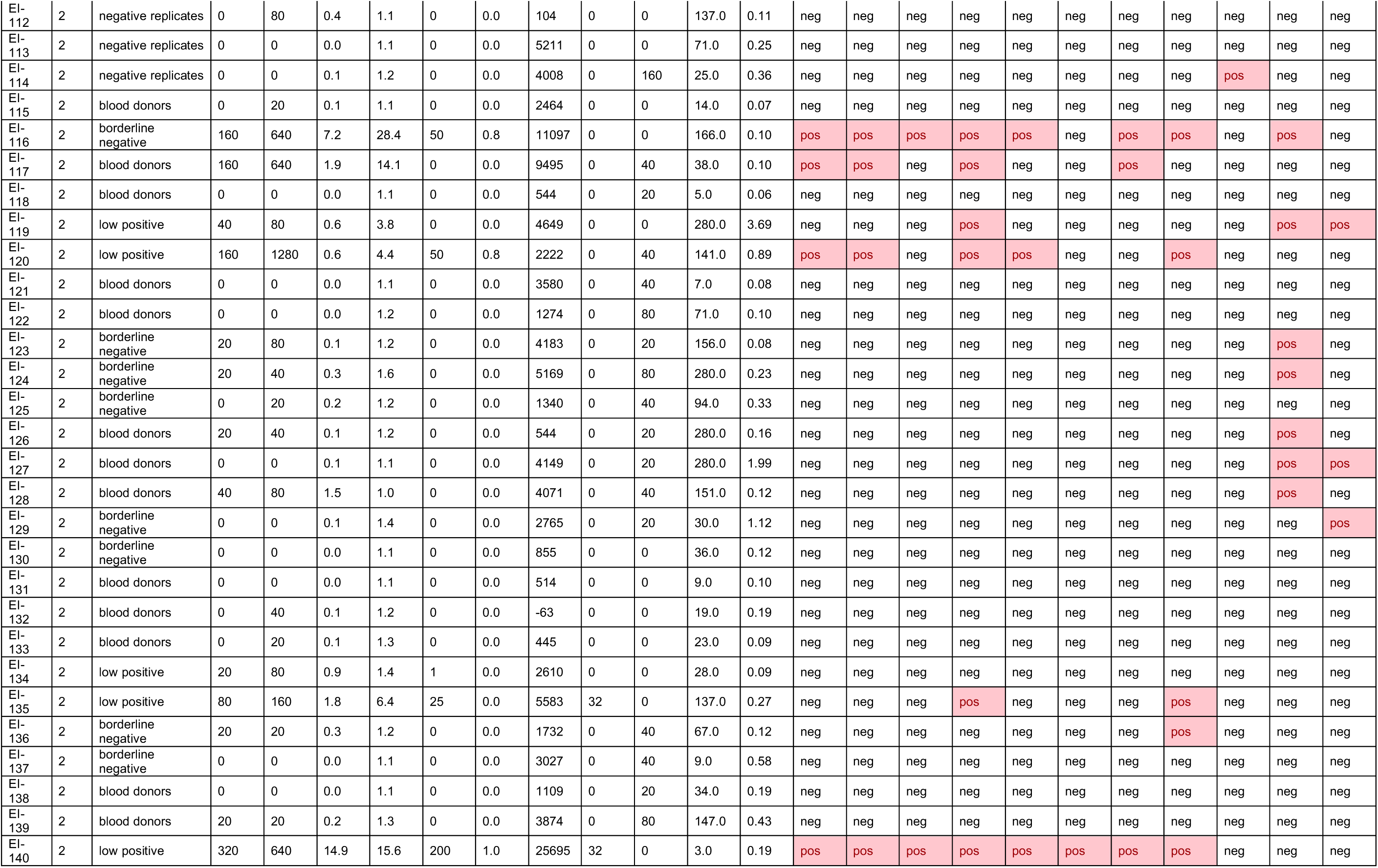
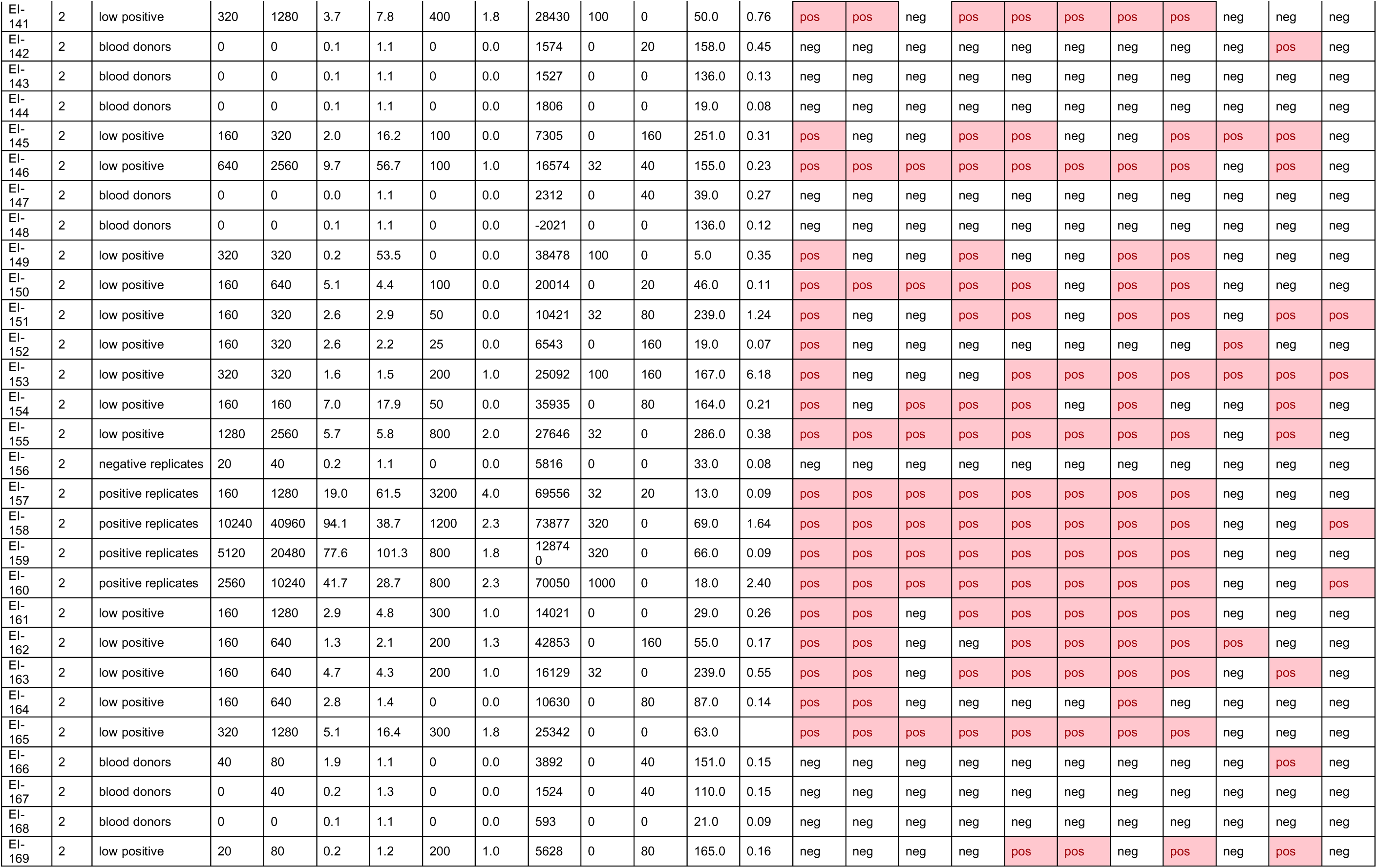
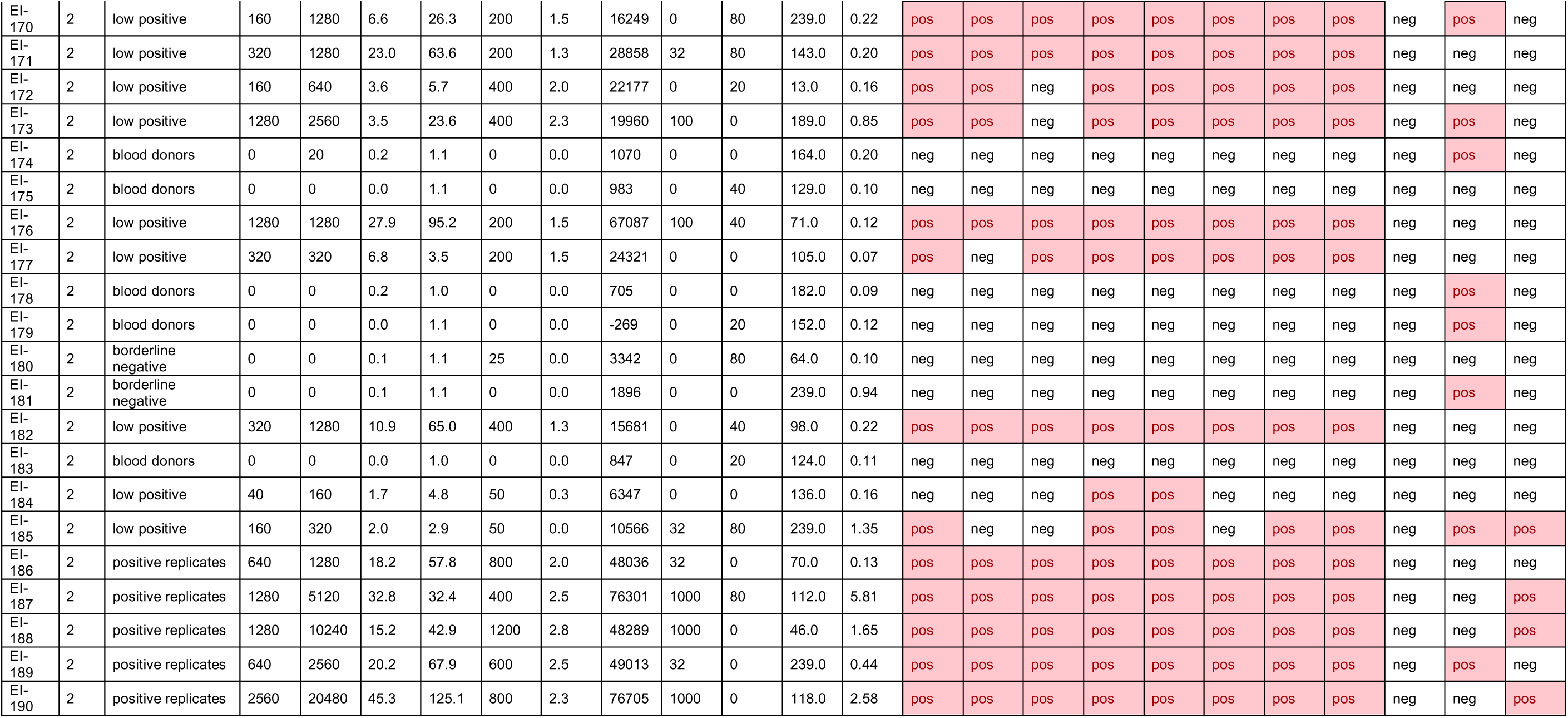
Raw data (qualitative and quanititative results) for all samples analyzed in this study according to the serostatus sent out for analysis.

